# Time to HIV rebound after antiretroviral therapy interruption: a double-blind randomised placebo-controlled trial of long-acting broadly neutralising antibodies; The RIO Trial

**DOI:** 10.64898/2026.02.04.25342277

**Authors:** Ming Jie Lee, Louise-Rae Cherrill, Panagiota Zacharopoulou, Simon Collins, Marcilio Fumagalli, Emanuela Falaschetti, Mohammed Altaf, Timothy Tipoe, Piyumika Godakandaarachi, Julie Fox, Alison Uriel, Amanda Clarke, Sabine Kinloch-de Loes, Sarah Pett, Marta Boffito, Gary Whitlock, Ole Schmeltz Søgaard, Kyle Ring, Irvine Mangawa, Jesal Gohil, Tamara Elliott, Henrik Nielsen, Jesper Damsgaard Gunst, Chloe Orkin, Rebecca Sutherland, Lisa Hamzah, Paola Cicconi, Graham P. Taylor, Jacquie Ujetz, Ishrat Jahan, Helen Brown, Nicola Robinson, Stephen Fletcher, Hanna Box, Kelly E. Seaton, Georgia Tomaras, Margaret E. Ackerman, Joshua A. Weiner, Anna Kaczynska, Cintia Bittar, Jill Horowitz, Michel Claudio Nussenzweig, Marina Caskey, John Frater, Sarah Fidler, the RIO Study Team

**Affiliations:** Imperial College London (M.J.L., L-R.C., E.F., G. Taylor., J.U., I.J., S.F., H. B., T.E., G.T., S. Fidler.); National Institute for Health Research (NIHR) Imperial Biomedical Research Centre (BRC) (S. Fidler., T.E., H. B.,); Nuffield Department of Medicine, the University of Oxford (P.Z., T.T., M.A., H. Brown.,N.R, P.C., J.F.); National Institute for Health Research (NIHR) Oxford Biomedical Research Centre (BRC) (J. F.); Department of Medicine and Therapeutics, Faculty of Medicine, The Chinese University of Hong Kong (T.T.); HIV i-Base, London (S.C.); Centre for Clinical Vaccinology and Tropical Medicine and the Oxford University Hospitals NHS Foundation Trust (P.C.); Guy’s and St Thomas Hospital NHS Foundation Trust (P.G., J. Fox.); Manchester University NHS Foundation Trust, Manchester (A.U., I.M.); University Hospitals Sussex NHS Foundation Trust and Brighton & Sussex Medical School (A.C.); Royal Free London NHS Foundation Trust (S.K-L.); University College London (S.K-L., S.P); Central and Northwest London NHS Foundation Trust (S.P.); St George’s University Hospital, London NHS Foundation Trust (L.H); Western General Hospital, NHS Lothian Trust, Edinburgh (R.S.); Chelsea and Westminster Hospital, London (M.B., G.W.); Queen Mary University of London and Barts Health NHS Trust, London (C.O., K.R.); Imperial College Healthcare NHS Trust, London (J.G.) – all in the United Kingdom; Aalborg University and Aalborg University Hospital, Aalborg, Denmark (H.N.); Aarhus University and Aarhus University Hospital, Aarhus, Denmark (O.S.S., J.D.G.); The Rockefeller University, New York, USA (M.F., C.B., A.K., J.H., M.C.N., M.C.); Howard Hughes Medical Institute, Chevy Chase, USA (M.C.N.); Duke University, Durham, USA (K.F.S., H.M.G, G.D.T); Dartmouth College, New Hampshire, USA (M.E.A., J.A.W.)

**Keywords:** HIV, cure, broadly neutralising antibodies, bNAbs, analytical treatment interruption, 10-1074, 3BNC117

## Abstract

**Background:** HIV-specific broadly neutralising antibodies (bNAbs) can maintain viral control after interrupting antiretroviral therapy (ART). We investigated the duration and efficacy of Fc-engineered long-acting bNAbs (LS-bNAbs) in maintaining ART-free HIV control compared with placebo.

**Methods:** RIO is a 1:1 randomised double-blind placebo-controlled trial of two LS-bNAbs (3BNC117-LS & 10-1074-LS) in individuals virally suppressed on ART initiated since early-stage HIV. Eligible participants interrupted ART after receiving blinded infusions of either both LS-bNAbs (Arm-A) or placebo (Arm-B). A second optional blinded infusion was offered after 20 weeks for participants who remained virally suppressed without ART. The primary outcome was time to viral rebound 20 weeks after ART interruption, defined as either the first of six consecutive plasma HIV RNA >1,000 copies/mL, or two measurements >100,000 copies/mL. Secondary outcomes included adverse events, long-term viral control and bNAb pharmacokinetics.

**Findings:** Sixty-eight participants were randomised, thirty-four to each arm. By week 20, viral rebound had occurred in 8 Arm-A and 30 Arm-B participants; 75% (95% CI, 61 – 92) of Arm-A participants had not rebounded, compared to 11% (95% CI, 4-29) participants in Arm-B. Arm-A participants were 91% less likely to rebound compared to Arm-B participants (hazard ratio of rebound (HR): 0.09; 95% confidence interval (CI) 0.04 – 0.21, P < 0.001). ART-free viral control using the above endpoints beyond 96 weeks was evident in 7 (25%, 95% CI 13 – 48) Arm-A participants compared to 2 (11%, 95% CI 4 – 29) Arm-B participants (HR: 0.22, 95% CI 0.12 – 0.40, P < 0.001). These seven participants had predicted serum bNAb concentrations below the presumed therapeutic threshold of 10 μg/mL at 96 weeks. Of nine serious adverse events, none were study-related.

**Conclusion:** Long-acting bNAbs can sustain extended ART-free viral control in people treated during early-stage HIV and represent a promising step towards achieving ART-free HIV remission.

**Funding:** The RIO trial was funded by the Bill & Melinda Gates Foundation (grant ref. OPP1210792). Infrastructure support in the UK for this research was provided by the National Institute of Health Research (NIHR) Imperial Biomedical Research Centre (BRC), the NIHR Imperial Clinical Research Facility (ICRF) and the Oxford NIHR BRC.

- Study/trial registration numbers and date of registration:
  ○ UK Research Ethics Committee reference: 19/LO/1669 (11 Sep 2019)
  ○ EudraCT: 2019-002129-31 (12 Dec 2019)
  ○ EU CTR: 2024-514564-13-00 (02 Jan 2025)
  ○ ClinicalTrials.gov Identifier: NCT04319367 (02 Mar 2020)
  ○ UK IRAS: 266322
  ○ Sponsor Protocol Number: 19IC5249
  ○ Funder Reference: OPP1210792

**Evidence in context:** We systematically searched Medline, Embase, and Web of Science databases until April 2024 with additional searches updated until January 2026. Combination therapy of HIV-specific broadly neutralising antibodies (bNAbs) has demonstrated periods of viral control for people living with HIV who interrupt their antiretroviral treatment. The development of long-lasting LS-bNAbs with Fc-receptor modifications has been shown to increase the serum half-lives by up to four-fold. To date, the clinical efficacy and duration of ART-free HIV control with LS-bNAbs has not been evaluated in a sufficiently powered human trial.

**Added value of this study:** The RIO trial; a prospective double blind randomised controlled study for the first time demonstrates that a single dose of two bNAbs 10-1074-LS and 3BNC117-LS was 91% more effective in maintaining ART-free viral control to twenty weeks compared to placebo. The long-term follow up to 96 weeks adds new data on the duration and frequency (25%) of viral control conferred by two doses of LS-bNAbs beyond the expected frequencies of post-treatment control in an early treated cohort of people living HIV. ART-free viral control beyond 96 weeks is likely due to post-bNAb induced mechanisms as modelled bNAb concentrations past this time were below presumed therapeutic thresholds. Administration of LS-bNAbs was safe and not associated with any Severe Adverse Events.

**Implications of all the available evidence:** LS-bNAbs represent a major advance towards ART-free HIV control and remission as an achievable goal. These findings will inform future combination strategies to enhance the mechanisms of post-bNAb HIV control.

## Introduction

Antiretroviral therapy (ART) has transformed survival for people living with HIV and prevents viral transmission^1^, but alone cannot cure HIV due to the persistence of a reservoir of latently infected cells^2^. As a result, ART is a lifelong therapy, introducing challenges of sustaining global access, funding, stigma, drug-related side effects^3^, and treatment fatigue^4,5^. There is an enduring drive and need for safe and effective long-lasting therapies aimed at achieving either a cure or long-term ART-free remission.

ART blocks viral replication, maintaining plasma viral suppression below the limit of detection. ART interruption is usually followed by a rapid viral rebound within 2-4 weeks^6^. The HIV reservoir is the source of the rebounding virus^7^. The absence of assays to accurately predict viral rebound means the only way to determine efficacy of novel interventions is to interrupt ART; a process called analytical treatment interruption (ATI) ^8^.

HIV-specific broadly neutralising antibodies (bNAbs) targeting conserved regions on the HIV envelope glycoprotein neutralise a broad range of HIV strains, maintaining viral control after ART is interrupted^9^. Single bNAb studies including an ATI demonstrated a variable but delayed time to viral rebound when dosed either on ART^10,11^ or at ART initiation^12,13^. When used as monotherapies in viraemic people with HIV, following an initial periods of decreased viraemia, the presence of bNAbs select for bNAb-resistant strains^10,11^. However, when used in combination, bNAbs 3BNC117 (anti-CD4-binding site)^14^ and 10-1074 (anti-V3 loop),^15^ conferred much longer periods of viral control in the absence of ART^16–18^. Engineering the Fc portion of a bNAb extends their half-life through modification of the interactions with the neonatal Fc receptor (FcRn). Mutations such as M428L/N434S (LS) at the Fc–FcRn interaction enhance FcRn binding and intracellular recycling, increasing antibody half-life^19^. This ‘LS’ modification extends bNAb half-lives 3-4-fold e.g. from 24 to 81 days for 10-1074-LS^19,20^. To date, the clinical efficacy and duration of ART-free viral control with long-acting bNAbs has not been evaluated in a sufficiently powered human trial.

The RIO study is an ongoing double-blind randomised placebo-controlled trial comparing the safety, duration and efficacy of two LS-bNAbs, 3BNC117-LS and 10-1074-LS, with placebo, on time to viral rebound following ART interruption in people with HIV treated with ART since early-stage infection.

## Methods

### Study design

RIO is a two stage placebo-controlled double-blind two-arm prospective phase II randomised controlled trial. The RIO study protocol has been previously published ^21^ and is included in the supplementary materials. Further details of the risk mitigation steps, for the monitoring of participants undertaking a treatment interruption study, COVID-19 pandemic prevention measures, and prevention of HIV transmission through ensuring access of HIV pre-exposure prophylaxis (PrEP) for partners are included in the published protocol ^21^.

### Eligibility Requirements

People living with HIV were eligible if they initiated ART during confirmed primary or early-stage HIV^21^, with the latter defined as a nadir CD4 count >500 cells/µL. Participants were screened for predicted sensitivity to 10-1074 using proviral single-genome amplification of *env* genes and an *in silico* prediction algorithm, as previously described ^18,22^ (see Supplementary Methods).

### Randomisation and Treatments

Randomisation was performed centrally using a web-based system. Block randomisation with variable block sizes of 2 or 4 was used, without stratification. Participants, care providers, and outcome assessors were blinded to the assignment. In stage 1 of the protocol, participants were assigned 1:1 to receive one infusion of 10-1074-LS (10 mg/kg) and 3BNC117-LS (30 mg/kg) (Arm-A), or saline placebo infusion (Arm-B). Two days post-infusion, ART was interrupted. Following early data indicating that multiple doses of bNAbs were associated with relative changes in the intact proviral reservoir over six months^18^, a second-dose sub-study (protocol amendment approved on the 22^nd^ June 2022) was introduced to allow an optional second blinded infusion was offered after 20 weeks if viral rebound had not occurred.

Participants were unblinded and resumed ART once they met the protocol-defined criteria for viral rebound. In response to feedback from clinicians, participants, and the community, a subsequent protocol amendment allowed participants to provide consent for unblinding after 48 weeks while remaining off ART, extending the duration of follow-up data collection (Supplementary Protocol). Participants who were unblinded to placebo were offered open-label LS-bNAbs in the second stage of the protocol. Only data from the first stage are reported here, as the second stage of RIO is ongoing.

### Assessments

During ATI, weekly clinical assessments, CD4 counts, and plasma HIV viral load (VL) measurements were performed for 8 weeks. Assessments were then conducted fortnightly until week 20, and every four weeks thereafter until viral rebound. Once VL became detectable, assessments returned to a weekly schedule. VLs were measured using validated assays at local clinical sites, with undetectable results defined as <50 copies/mL (see Table S1). Serial serum bNAb concentrations were measured by a validated anti-idiotype ELISA^16,23^. Anti-drug antibodies (ADA) were assayed at pre-and post-bNAb infusion timepoints using a three-tiered electrochemiluminescence approach described by Bharadwaj et al.^24^, longitudinally up to six months after the last bNAb dose. (see Table S2). Plasma concentrations of tenofovir disoproxil, lamivudine, and dolutegravir were measured at multiple timepoints in participants with VL <50copies/mL using validated liquid chromatograph-mass spectrometry assays.

### Outcome Measures

Viral control was defined as the time from ART interruption to plasma viral rebound. The primary outcome, originally defined at 36 weeks, was reported at 20 weeks after treatment interruption in accordance with the protocol amendment, which offered participants a second dose after this timepoint. Rebound was defined as either two consecutive VL measurements >100,000 copies/mL, six consecutive weeks >1,000 copies/mL, or other protocol-defined instances including participant preference to restart ART^21^. Dates of rebound were confirmed by an independent Endpoint Adjudication Committee (EAC) blinded to study arm.

### Secondary outcomes

Secondary outcomes include time to rebound beyond 20 weeks, Adverse Events (AEs) and Serious Adverse Events (SAEs) by study arm, time to viral rebound by different VL thresholds, CD4 T cell counts and CD4:CD8 ratios during ATI. We also reported time to viral resuppression after restarting ART, longitudinal serum bNAb concentrations, and baseline predicted bNAb resistance. A full list of secondary and exploratory outcomes are included in the supplementary protocol. Secondary outcomes were initially designed to capture outcomes up to 48 weeks after ATI. However, the second-dose study extension enabled reporting of outcomes up to 96 weeks after ATI for participants still in follow-up at the time of database lock.

### Statistical Analysis

RIO is a parallel-group trial that uses a superiority framework. Median follow-up was calculated using the reverse Kaplan-Meier analysis method. Survival proportions were estimated using the Kaplan-Meier method. Time-to-event analyses for the primary and secondary outcomes were performed using an intention-to-treat approach. Differences in the time-to-viral rebound were assessed using the log-rank test and the Cox proportional-hazard model at 20 weeks and 96 weeks. Participants who restarted ART before reaching the primary endpoint or viral rebound were censored at the time of ART restart or last viral load measurement. All statistical results were two-tailed, with a 5% significance level. Safety outcomes were reported descriptively. Statistical analysis was performed using R version 4.5.0.

### Sample size calculation

The trial was designed to detect a reduction in the rebound rate at 36 weeks from 90% in Arm-B to 55% in Arm-A (hazard ratio of rebound (HR) 0.35). At 5% significance level and 90% power, 36 participants per arm were required, allowing for a 10% drop-out rate. The primary outcome timepoint was reduced from 36 to 20 weeks, due to the second-dose sub-study extension (see Supplementary Protocol and Statistical Analysis Plan). Statistical power was not compromised, as viral rebound rates in the placebo arm were predicted to remain at 90% (6).

### Viral dynamics

Plasma viral load natural log doubling time (DT) was analysed as an exploratory endpoint using measurements between and inclusive of the first detectable plasma HIV RNA measurement >50 copies/mL since ATI, and the first peak plasma HIV RNA >1000 copies/mL before ART restart. The slope of increase (r) was calculated by linear regression and viral doubling time (t) calculated using the equation

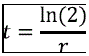

Participants who withdrew from the first ATI or those that did not experience viral rebound were excluded from this analysis.

### Pharmacokinetic analyses

Serum bNAb levels were measured using a validated anti-idiotype ELISA ^16,23^ where samples were available. Due to the well defined PK of these antibodies, these data were used to derive a two-compartment model with Monolix^TM^ software using the Stochastic Approximation for Model Building Algorithm ^25^ to infer missing values. The following covariates were tested: age, log(weight), HIV-1 clade, predicted baseline bNAb resistance mutations, and number of doses. The model that best fit the data for both bNAbs was a two-compartment model with a central compartment (V), a peripheral compartment with rates of transfer to and from represented by the constants k_12_ and k_21_, and a linear elimination with a rate constant k. Parameters were transformed into A, B, alpha and beta using pharmacokinetic equations listed below. Ninety-five percent prediction intervals (PI) were calculated using RsmLx in R with 1,000 bootstrap replicates.

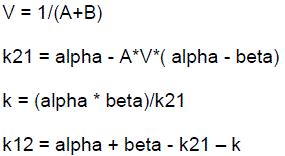

Pharmacokinetic simulations using Simulx were run with 100 replicates to estimate bNAb serum concentration estimates up to 1,000 days following the first infusion. Estimated rebound concentrations at the time of viral rebound were reported, however, due to delays in receiving serum samples for bNAb ELISA assays, not all timepoints from all participants were available. In addition, because the timing of viral rebound was determined retrospectively, samples at the exact time of rebound may not have been collected. A bNAb concentration threshold of 10 μg/mL was selected as in previous studies involving both bNAbs, viral rebound was not observed until at least one bNAb level had fallen below 10 μg/mL^16,18^. Additionally, animal studies have shown that serum concentrations required for *in vivo* protection (IC50) are typically one to two orders of magnitude higher than corresponding *in vitro* measurements ^26^.

### Ethical and regulatory governance

RIO is led by the Trial Management Group, which includes a representative from the community of people living with HIV. The study protocol received ethical approval from the London – Westminster Research Ethics Committee (REC reference: 19/LO/1669) on September 11, 2019. The Imperial Clinical Trials Unit provides oversight for the operational and regulatory conduct of the study. Independent Data Monitoring and Trial Steering Committees convenes biannually to review trial safety and overall conduct, respectively.

## Results

### Participants

Of the 145 people screened from May 2021 to July 2024 (Figure 1), 77 were ineligible, including 27 (18.6%) with predicted insensitivity to 10-1074. Sixty-eight participants were enrolled across nine UK and one Danish site. The cut-off date for the analysis presented in this publication was September 26, 2025. Median ATI follow-up time was 117 weeks (range 1 – 195 weeks). Table 1 summarises the 68 enrolled participants. Baseline characteristics were balanced between study arms. Median age was 40 years, 85% were of white ethnicity, all were cis-gender men, and 56% had HIV clade B virus. Median baseline CD4 count was 776 cells/μL (Interquartile (IQR 626 – 963). Most participants (91%) had started ART in primary HIV infection and received an integrase-inhibitor-based regimen at enrolment. Median duration on ART was 5 years (IQR 3-6).

**Figure 1:**
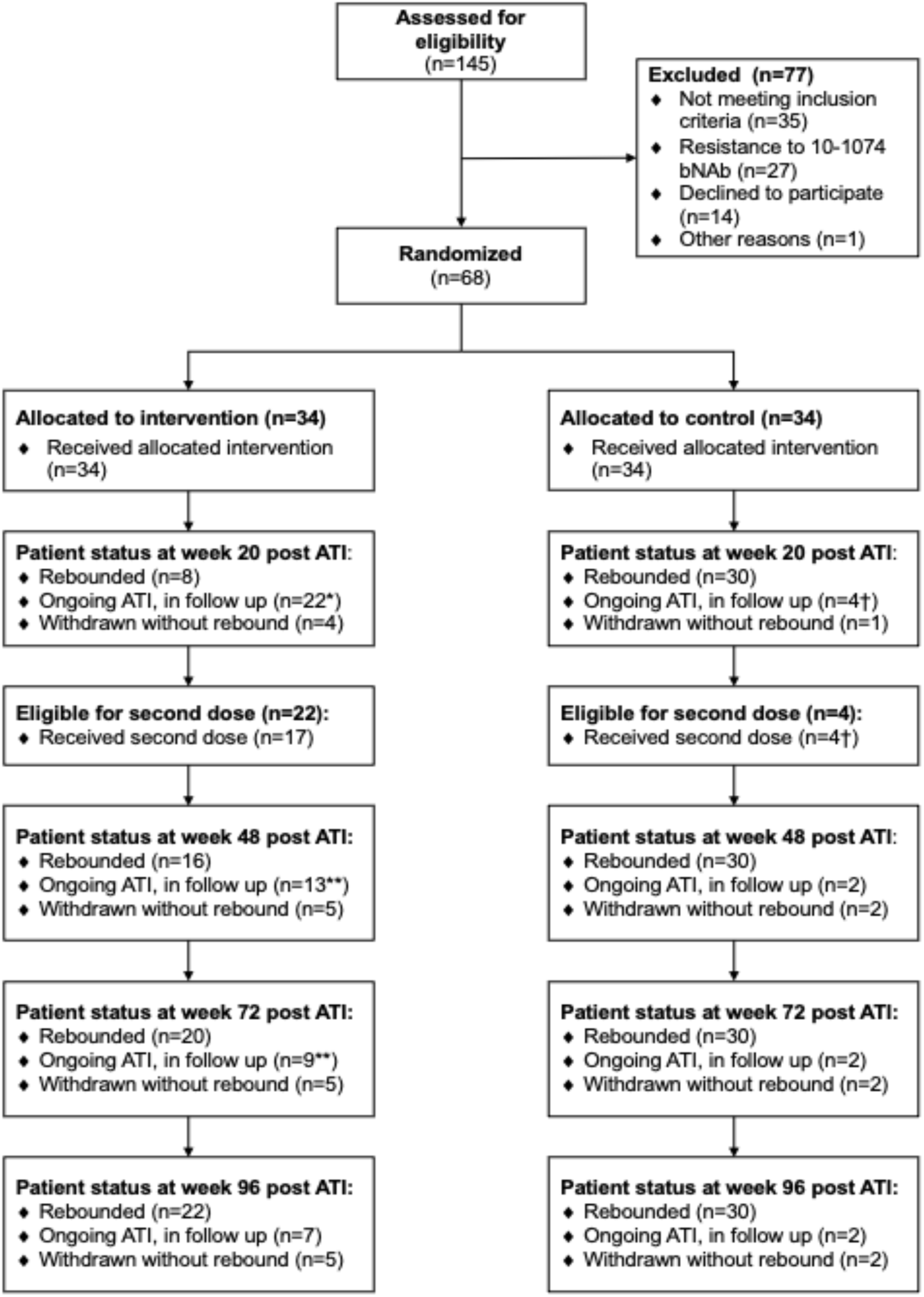
Study consort diagram. *Number includes one participant who temporarily restarted ART between week 12 – 20 of their ATI due to concurrent illness, and rebounded at week 31. **Number includes one participant who temporarily restarted ART between week 48 – 55 of their ATI due to concurrent illness, and rebounded at week 97. † Number includes one participant who had two doses of placebo infusions and continued their ATI until week 48. Time to rebound was retrospectively determined by the endpoint adjudication committee as 10 weeks, they were included in both rebound and ongoing ATI numbers.

**Table 1.** Participant demographics.

|  | **Arm-A: bNAbs** | **Arm-B: Placebo** | **All** |
| --- | --- | --- | --- |
|  | **(N=34)** | **(N=34)** | **(N=68)** |
| **Age, years** |  |  |  |
| Median (IQR) | 36 (31, 44) | 42 (33, 51) | 40 (32, 46) |
| **Sex assigned at birth, n (%)** |  |  |  |
| Male | 34 (100.0%) | 34 (100.0%) | 68 (100.0%) |
| **Gender identity, n (%)** |  |  |  |
| Cis-gender men | 34 (100.0%) | 34 (100.0%) | 34 (100.0%) |
| **Weight, kg** |  |  |  |
| Median (IQR) | 73 (70, 81) | 78 (71, 89) | 77 (70, 85) |
| **BMI, kg/m<sup>2</sup>** |  |  |  |
| Median (IQR) | 23 (22, 26) | 25 (23, 28) | 24 (22, 27) |
| **Ethnicity, n (%)** |  |  |  |
| Asian or Asian British | 3 (8.8%) | 0 (0.0%) | 3 (4.4%) |
| Black or Black British | 1 (2.9%) | 2 (5.9%) | 3 (4.4%) |
| Other | 2 (5.9%) | 2 (5.9%) | 4 (5.9%) |
| White | 28 (82.4%) | 30 (88.2%) | 58 (85.3%) |
| **Baseline CD4 count, cells/μL** |  |  |  |
| Median (IQR) | 742 (657, 937) | 810 (596, 986) | 776 (626, 963) |
| **HIV clade, n (%)** |  |  |  |
| B clade | 19 (76.0%) | 21 (84.0%) | 40 (80.0%) |
| Non-B clade | 6 (24.0%) | 4 (16.0%) | 10 (20.0%) |
|  | **(N=34)** | **(N=34)** | **(N=68)** |
| Information not available | 9 | 9 | 18 |
| **ART regimen at screening, n (%)** |  |  |  |
| NRTI | 34 (100.0%) | 33 (97.0%) | 67 (98.5%) |
| INSTI | 27 (79.4%) | 31 (91.2%) | 58 (85.3%) |
| bPI | 3 (8.8%) | 3 (8.8%) | 6 (8.8%) |
| **Duration on ART prior to enrolment, years** |  |  |  |
| Median (IQR) | 5 (3 – 6) | 5 (4 – 7) | 5 (4 – 7) |
| **Stage of HIV at time of diagnosis** |  |  |  |
| Primary HIV | 32 (94) | 30 (88) | 62 (91) |
| Early HIV* | 2 (6) | 4 (12) | 6 (9) |

IQR: Inter-quartile range, BMI: Body mass index, ART: Antiretroviral Therapy, NRTI: Nucleoside reverse transcriptase inhibitor, INSTI: Integrase strand transfer inhibitor, bPI: boosted protease inhibitor. \*Early HIV was defined as having been diagnosed and started ART while the nadir CD4 count was above 500 cells/ μL.

### Baseline bNAb screening

Of the 68 participants enrolled, 28/34 Arm-A and 30/34 Arm-B participants had proviral *env* sequences with no or <15% sequences containing resistance-associated mutations to 10-1074, meeting the screening criteria previously described^21^. Env sequences from six Arm-A and four Arm-B participants were not possible to amplify at baseline, and these participants were considered eligible. Two of these participants were subsequently found to harbour resistance-associated mutations to 10-1074 at post-hoc sequencing.

### Treatments

All participants received their randomised dosing. Four participants in Arm-A and one participant in Arm-B restarted ART before meeting the viral rebound criteria prior to week 20. One Arm-A participant died from a myocardial infarction before reaching week 20. This death was reviewed as unrelated to study drug or procedures. Of those maintaining viral control off ART beyond 20 weeks, two additional participants (one in Arm-A, one in Arm-B) chose to restart ART before viral rebound by 96 weeks. Of 26 eligible participants, 21 received a second infusion; the median time of the second blinded infusion was 27 weeks (range 21 – 50 weeks) after randomisation. Two participants temporarily resumed their ART during ATI due to concurrent illness and subsequently reinitiated treatment interruption and were included in the final analysis, in accordance with the protocol.

### Viral control within 20 weeks (Primary outcome)

By week 20, viral rebound had occurred in 8 Arm-A and 30 Arm-B participants. Seventy-five percent (95% CI, 61 – 92) of Arm-A participants had not rebounded, compared to 11% (95% CI, 4 – 29) of participants in Arm-B after censoring. In a Cox proportional-hazards regression, Arm-A participants were significantly less likely to rebound compared to Arm-B (HR 0.09; 95% CI, 0.04 – 0.21; P <0.001) (Figure 2a). The difference in restricted mean survival time was 11.9 weeks (95% CI, 9.7 – 14.0).

**Figure 2:**
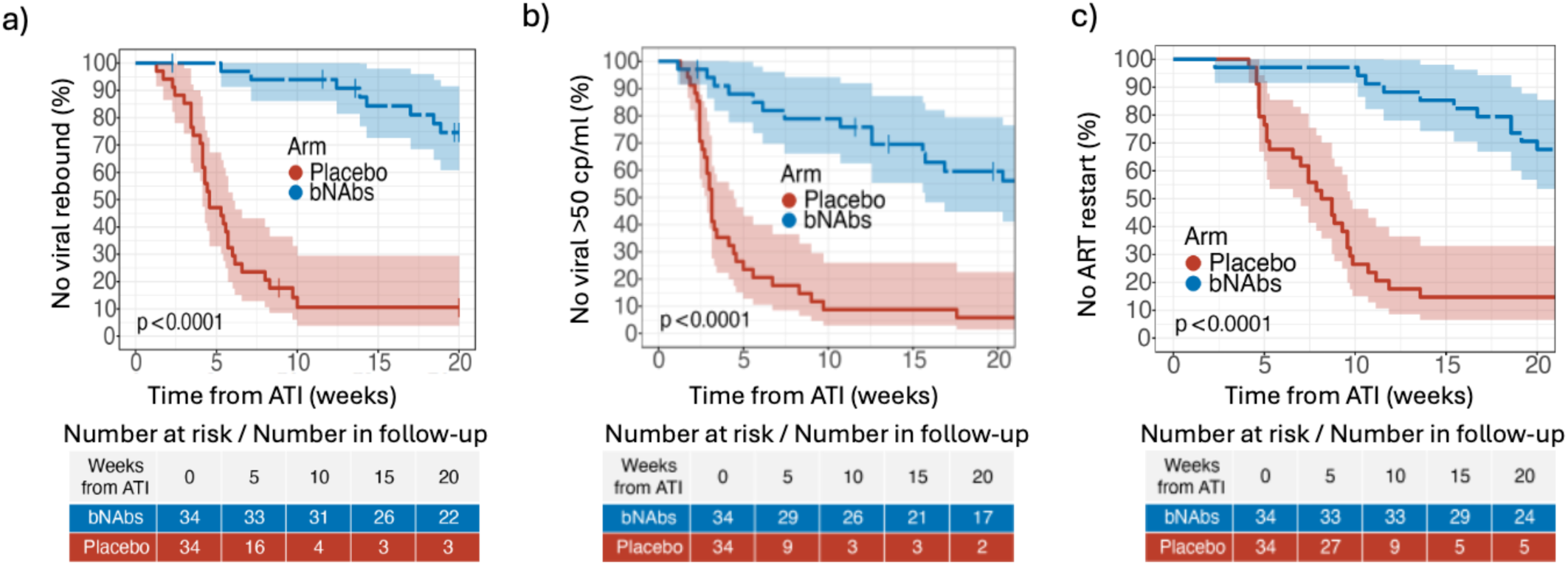
Endpoint outcomes by week 20. **a)** Kaplan-Meier (KM) survival curves showing the proportion of participants who had not experienced viral rebound by the week 20 primary endpoint viral rebound criteria during the stage 1 ATI of the RIO study, stratified by study arm **b)** KM survival curves showing the proportion with undetectable viral loads during the ATI to 20 weeks stratified by study arm. **c)** KM survival curves showing the proportion who have not restarted ART during the ATI to week 20. Shaded areas represent the 95% confidence intervals.

The results were consistent using different outcome thresholds at week 20. At week 20, the proportion of participants with undetectable viral loads (<50 copies/mL) was 60% (95% CI, 45 – 79) in Arm-A and 6% (95% CI, 2 – 23) in Arm-B (Figure 2b). By week 20, 68% (95% CI, 54 – 85) of Arm-A participants remained ART-free and 15% (95% CI, 7 – 33) in Arm-B (Figure 2c).

### Viral control beyond 20 weeks

ART-free viral control at 96 weeks post ART-interruption was evident in 25% (95% CI, 13 – 48) of Arm-A participants (n=7), compared to 11% (95% CI, 4 – 29) of Arm-B participants (n=2) (Figure 3a). Median time from randomisation to viral rebound was estimated to be 4.6 weeks (95% CI, 4.1 – 6.0) in Arm-B and 45.4 weeks (95% CI, 27.9 – 90) in Arm-A. In a Cox proportional-hazards regression, Arm-A participants were significantly less likely to rebound compared to Arm-B (HR 0.22; 95% CI, 0.12 – 0.40; P <0.001). Median time to rebound after last bNAb dose in Arm-A was 31.1 weeks (95% CI, 23.3 – 68.0).

**Figure 3:**
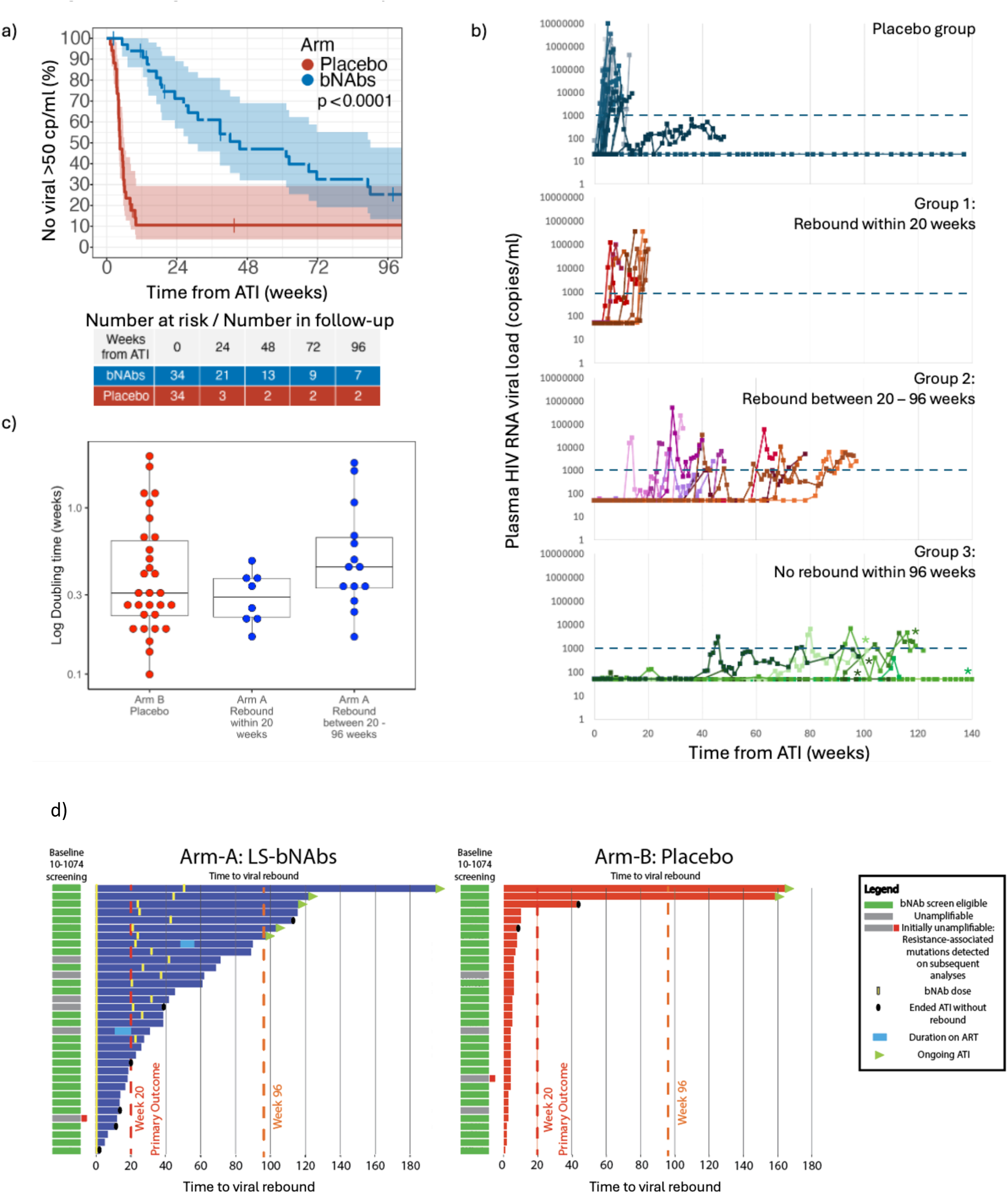
Longer term outcomes beyond week 20. **a)** Kaplan-Meier (KM) survival curves showing the proportion of participants who had not experienced viral rebound by the week, stratified by study arm. Shaded areas represent the 95% confidence intervals. **b)** Patterns of viral rebound of individual participants in both placebo and intervention arms plasma HIV RNA levels, defined by time of rebound in the interventional arm: group 1 – Early rebound within 20 weeks of ATI (n=8), group 2 – Rebound between 20 – 96 weeks (n=14), and group 3 – No rebound within 96 weeks (n=7). * denotes participants who remain off ART at the time of writing. The dashed lines represent a plasma HIV RNA viral load threshold of 1000copies/mL. **c)** Dot plot comparison of initial natural log viral doubling time in plasma HIV RNA viral load, between participants who received placebo infusions and participants who received bNAbs (arm-A), stratified by time of viral rebound by 20 (n=8) and 96 weeks (n=14) and placebo arm (n=30). **d)** Descriptive graphic representation of the relationship between the predicted baseline 10-1074 bNAb resistance and time since ATI start to viral rebound stratified by study arm. Each row represents individual participants. Screening for predicted 10-1074 eligibility are colour coded as per the legend. Participants who still remain off ART in an ongoing ATI are highlighted with green triangles. One of the unamplifiable Arm-A samples and one in Arm-B at baseline were subsequently assessed to be fully resistant after the participant was enrolled. Week 20 (Primary Outcome) and week 96 timepoints are represented by the red and orange dashed lines respectively.

For Arm-A participants, we observed three viral rebound patterns by time from ATI (Figure 3b). First, eight participants rebounded within 20 weeks. Second, 14 participants rebounded between 20 to 96 weeks with predicted measured serum concentrations of bNAbs above the assumed therapeutic threshold of 10ug/mL at the time of viral rebound. Third, seven participants remained off ART at and beyond 96 weeks, with predicted serum bNAb concentrations below 10ug/mL. Five of these participants still remain off ART between 98 – 195 weeks with their ATI ongoing. One participant chose to restart ART at week 113 without viral rebound, and one experienced viral rebound at week 116. These seven participants all had a second bNAb dose at a median time of 25 weeks after the first dose (range 21 – 50). Of note, no ART drugs were detectable in the plasma in any participant undetectable during the ATI.

To examine if LS-bNAbs treatment altered subsequent viral rebound kinetics, we compared viral rebound doubling times (DT) between study arms. HIV rebound DT was evaluated across groups experiencing viral rebound by study arm. Participants who rebounded within 20 weeks after receiving LS-bNAbs in Arm-A demonstrated doubling times (median DT 0.29 weeks IQR 0.22 – 0.38) comparable to placebo (0.31 weeks IQR 0.22 – 0.64). Those who rebounded after 20 weeks showed a median DT of 0.44 (0.33 – 0.66) weeks (Figure 3c). As these post-hoc analyses were exploratory in nature no formal statistical comparison was undertaken. Participants who did not have amplifiable baseline sequences were not associated with different viral rebound outcomes to those with amplified sequences (Figure 3d).

### Viral rebound dynamics

Median peak HIV RNA viral load at rebound was also lower in 17 participants who received 2 doses of LS-bNAbs compared to placebo (Arm-A, 2 doses: 6,880 copies/mL (range 1,280 – 58,700) vs Arm-B: 97,750 copies/mL (range 3,236 – 10,000,000)).

### LS-BNAb pharmacokinetics

Serum LS-bNAbs were detected in available samples from 31 Arm-A participants and fitted to a two-compartment model (Figures 4a-b). The estimated elimination half-lives of 10-1074-LS and 3BNC117-LS were 72.5 days (95% PI 66.0 – 78.8 days), and 64.7 days (95% PI 60.8 – 69.1days), respectively (see Table S4). Fifteen of seventeen participants who had two LS-bNAb doses had measurements available after the second dose. Variable times of LS-bNAb second dose are represented in Figure 4c. No anti-drug antibodies (ADAs) were detected in post-infusion samples from 11 Arm-A participants tested up to six months post-bNAbs (Table S2). No ART drug concentrations were detected in plasma samples.

**Figure 4:**
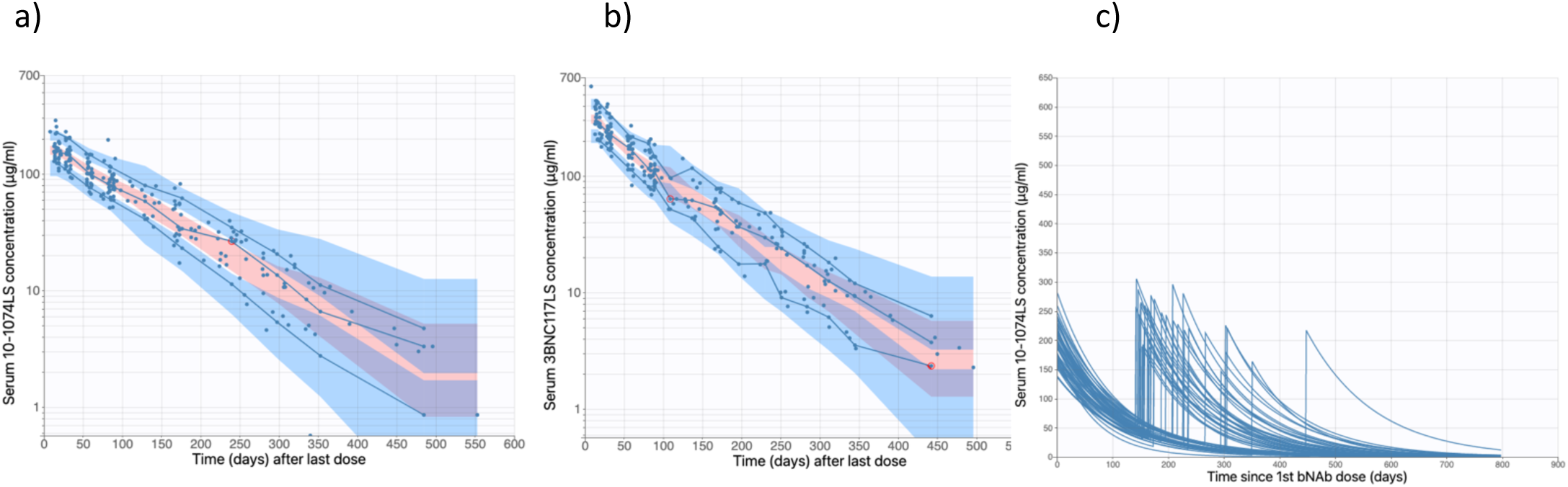
Pharmacokinetics of bNAbs. **a)** 10-1074-LS and **b)** 3BNC117-LS serum bNAb measurements are shown fitting a two-compartment pharmacokinetic model in these visual predictive charts, with serum bNAb concentrations (μg/mL) on the y-axis and time from the last dose (days) on the x-axis. Each dot represents a single bNAb measurement, and the lines represent the 10^th^, 50^th^ and 90^th^ percentiles respectively. The shaded areas represent the 90% prediction intervals. **c)** Serum 10-1074-LS concentrations are modelled out to 800 days after randomisation, to demonstrate the range of 2^nd^ dose times and peak serum concentrations after two doses.

The individual LS-bNAb pharmacokinetics, plasma HIV RNA, and modelled LS-bNAb concentrations at the time of viral rebound for all study participants are presented in Figures 5a-c. The median estimated concentration at rebound for 10-1074-LS and 3BNC117-LS was 40.6 µg/mL (range 0.6 – 149 μg/mL) and 52.4 µg/mL (range 0.4 – 231.4 μg/mL), respectively. Modelled population pharmacokinetic parameters and 95% prediction intervals are presented in Table S3.

Simulations were used to estimate the time for LS-bNAb concentrations to reach the previously assumed therapeutic threshold of 10μg/mL after two doses given 21 weeks apart. For 10-1074-LS and 3BNC117-LS, this was 47.6 weeks (95% PI 32.7 – 62.5) and 47.8 week (95% PI 35.6– 60.0) after the second dose, respectively.

**Figure 5:**
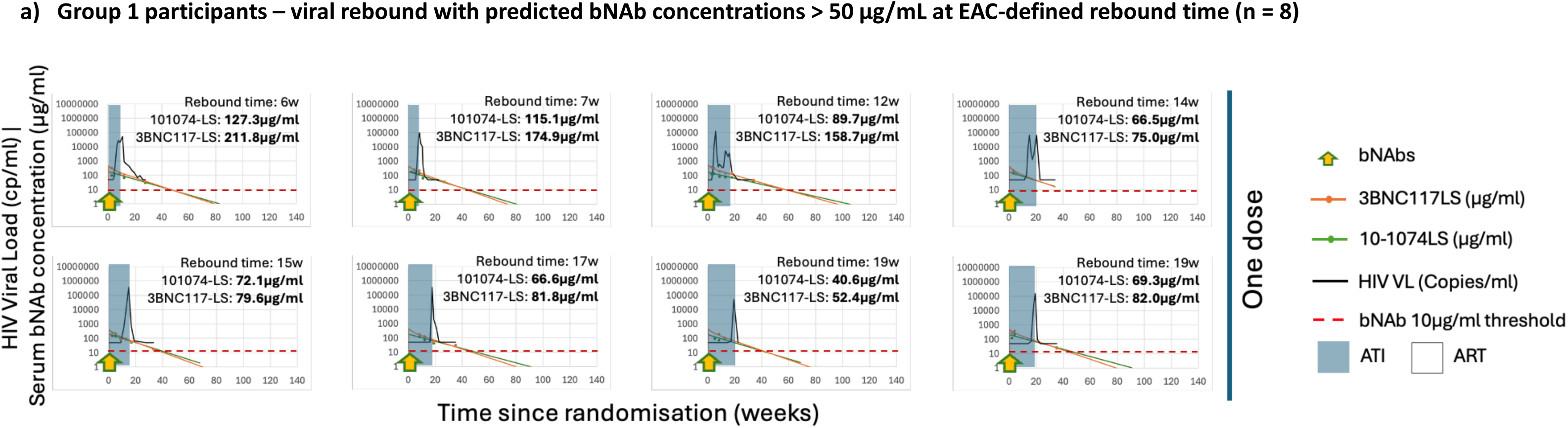
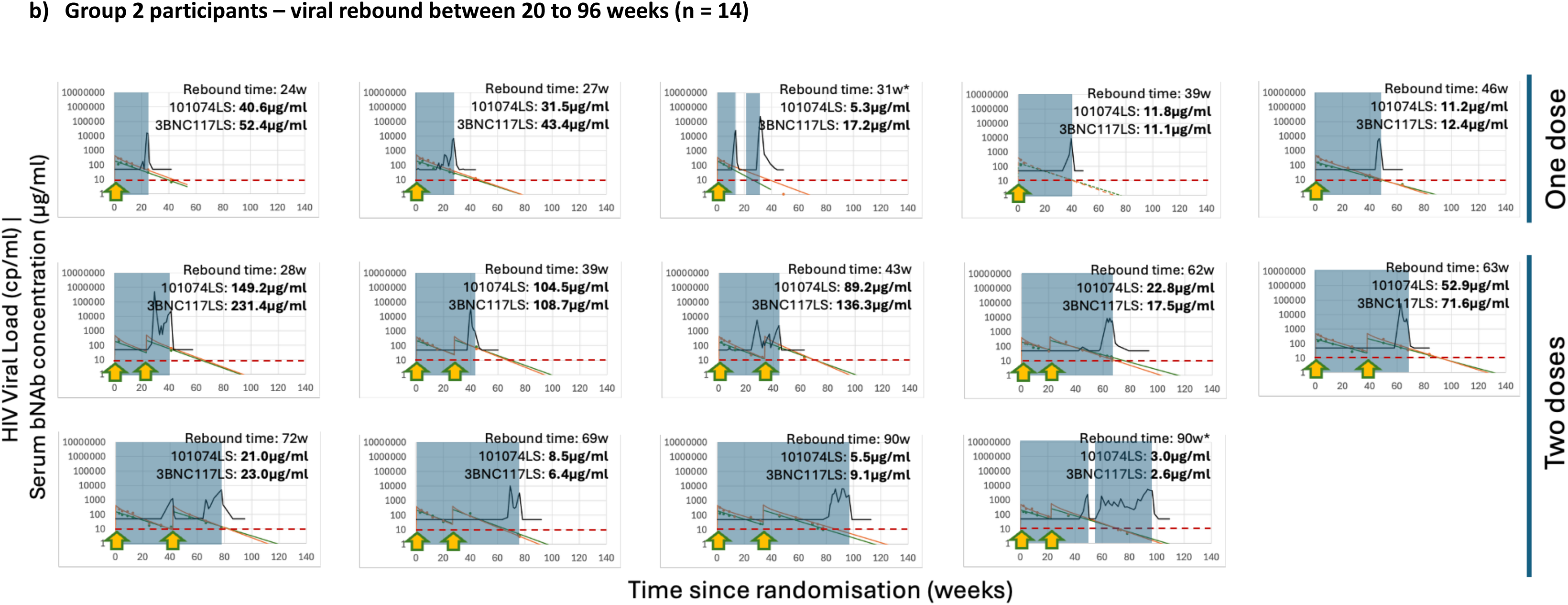
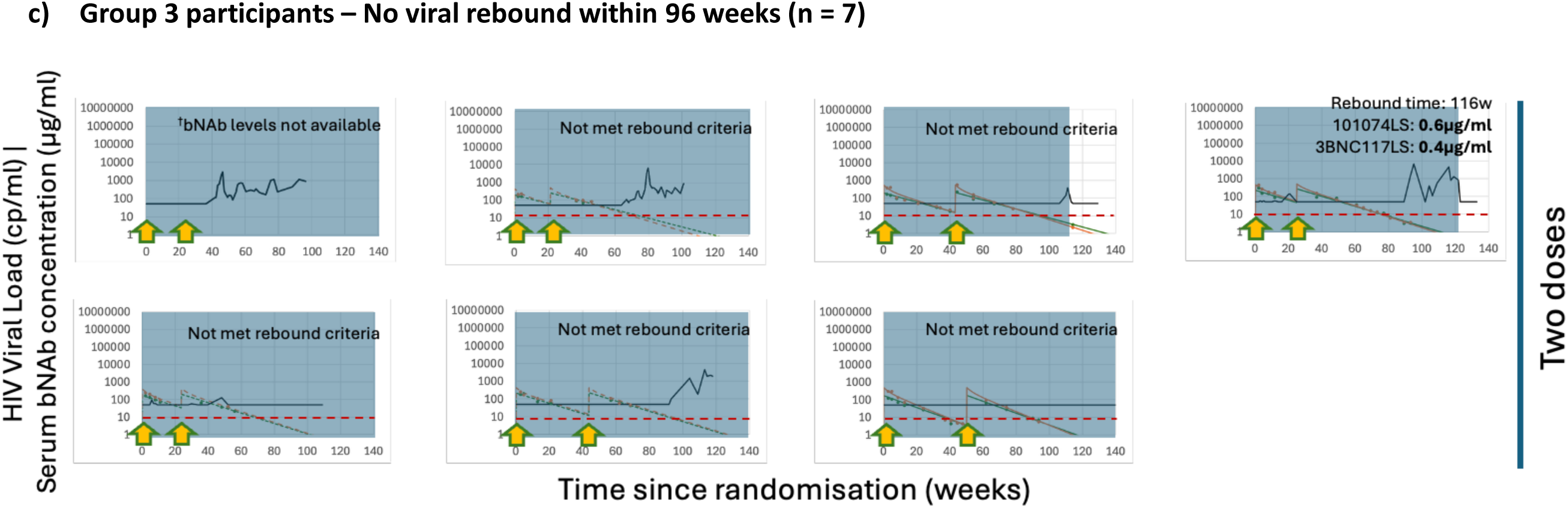
Relationship between viral loads and bNAb serum concentrations. Individual participants charts plotting their plasma HIV RNA viral load (black lines) and serum bNAb concentrations (green for 10-1074-LS and orange for 3BNC117-LS) against the time since randomisation in weeks on the x-axis, divided by modelled bNAb thresholds at EAC-defined time of rebound.. BNAb dosing times are indicated with arrows. The analytical treatment interruption (ATI) duration is shaded, and the time back on antiretroviral therapy (ART) in white. A threshold of 10μg/mL serum bNAb concentration is highlighted in the dashed red line. *These participants temporarily restarted ART (unshaded area) due to concurrent illness and interrupted treatment again once recovered in accordance with the study protocol. ^†^bNAb levels were not available for this participant. 16 arm-A participants had their time of rebound defined by the endpoint adjudication committee and did not meet either virological endpoints of 6 readings >1000copies/ml or 2 consecutive readings >100 000copies/ml. Participants who restarted ART or died before meeting ART restart criteria: n = 5

### Modelled bNAb concentrations at time of rebound

In the first group who rebounded within twenty weeks, the median 10-1074-LS concentration at time of rebound was 70.7 (IQR 66.6 – 96.0) μg/mL; median rebound 3BNC117-LS concentration was 82.0 (IQR 78.4 – 162.7) μg/mL) (Figure 5a). Participants who rebounded between 20 to 96 weeks had a median 10-1074-LS concentrations at time of rebound of 21.9 (IQR 9.2 – 47.5) μg/mL and median rebound 3BNC117-LS concentration of 20.3 (IQR 11.4 – 65.4) μg/mL (Figure 5b). All six participants with available bNAb measurements who remained off ART at 96 weeks had bNAb rebound concentrations below 10 μg/mL (Figure 5c). One participant did not have serum bNAb measurements available at the time of analysis, but remained off ART at 96 weeks where bNAb concentrations are expected to be below 10 μg/mL from population estimates (Figure 5c).

### Adverse Events

Thirty-two AEs were recorded as definitely or probably related to treatment or protocol (Table 2). Twelve SAEs have been recorded, none related to treatment or protocol. This included one death caused by a myocardial infarction associated with pre-existing undiagnosed triple-vessel cardiovascular disease (details summarised in Table S2).

**Table 2.** Adverse events.

|  | **Arm A (LS-**<br>**BNAb)** | **Arm B Stage 1**<br>**(Placebo)** | **Arm B Stage 2 (LS-**<br>**BNAb)** |
| --- | --- | --- | --- |
| **Total number of AEs** | 327 | 261 | 254 |
| **AE relation to study**<br>**procedure** |  |  |  |
| Definitely | 4 | 2 | 4 |
| Probably | 0 | 11 | 3 |
| Possibly | 11 | 17 | 1 |
| Unlikely | 39 | 43 | 35 |
| Not Related | 272 | 187 | 211 |
| **AE relation to Study IMP** |  |  |  |
| Definitely | 0 | 0 | 3 |
| Probably | 0 | 9 | 1 |
| Possibly | 10 | 15 | 2 |
| Unlikely | 43 | 46 | 34 |
| Not Related | 272 | 190 | 214 |
| Not assessable | 1 | 0 | 0 |
| **AE by severity** |  |  |  |
| Fatal (death) | 1 | 0 | 0 |
| Severe | 3 | 7 | 1 |
| Moderate | 46 | 54 | 15 |
| Mild | 276 | 199 | 239 |

### Time to viral resuppression and CD4 counts

Ninety-four percent of participants re-suppressed their plasma VL to undetectable levels within 12 weeks of restarting ART. There were no new ART drug resistance-associated mutations. All participants achieved viral re-suppression eventually. The median time to viral suppression after restarting ART for Arm-A was five (IQR 2 – 8) weeks, and three (IQR 2 – 5) weeks for Arm-B (Figure S1). Five Arm-B participants rebounded with peak VLs above 1,000,000 copies/mL (all asymptomatic), taking a median of five (range 2 – 24) weeks to achieve viral suppression, compared with none in Arm-A. There was no change in CD4 count or CD4:CD8 ratio during the period off ART in either study arm, and no HIV transmission events were recorded (see Table S4).

## Discussion

This is the first report of the extended duration and efficacy of two long-acting bNAbs on ART-free viral control amongst people starting ART in early-stage HIV compared to placebo. In the absence of ART, two HIV-specific LS-bNAbs reduced the risk of viral rebound within 20 weeks by 91% compared with placebo, following ART interruption. LS-bNAbs and ATI were not associated with adverse events. As well as confirming the efficacy of Fc-engineered LS-bNAbs at likely therapeutic concentrations, we demonstrate for the first time extended viral control when levels of serum bNAbs fall below this concentration. We report a significant increase in the frequency of participants who maintain viral control beyond 96 weeks, compared to expected frequencies of post-treatment control from both previous reports in the literature^6^, as well as in comparison with the placebo arm.

Previous studies with multiple doses of shorter-acting bNAbs^16,18^ demonstrated sustained viral control off ART for up to 48 weeks and observed viral rebound when 3BNC117 fell below 10 μg/mL. The TITAN study ^27^ demonstrated two doses of non-LS 10-1074 and 3BNC117, given 3 weeks apart with or without a TLR9-agonist, delayed viral rebound (median of 12.5 weeks after ATI), with no added benefit of the TLR9 agonist. Four of eleven (36%) of TITAN participants who received non-LS bNAbs maintained virologic control during the 25-week ATI, fewer than observed in RIO (75% by 20 weeks) following a single dose of the LS-variants. A consideration for RIO is that 17 participants received one bNAb dose and 17 received two. Clearly two doses will extend the duration of likely viral control, and this has been incorporated into the analyses. However, using measured and modelled PK data, it is still possible to estimate when sub-therapeutic bNAb levels are likely to have been achieved to allow analysis of potential post-bNAb control.

Studies with extended ATI periods highlight the importance of the developing standardised strategies to guide the safe management of participants, including the frequency of clinic visits, ensuring access to HIV PrEP for partners, and considering the practical, quality-of-life and psychological aspects of participation. RIO includes an ongoing qualitative sub-study to capture participant experiences as a key outcome, to inform the design of future ATI trials. The asymptomatic and unexpectedly high viral rebounds observed in some placebo recipients (> 1 million copies HIV RNA/mL) should also be noted. While ART-mediated viral resuppresion was rapid and effective in all study participants, the increased potential risk of transmission associated with such high viral rebounds must be highlighted in participant information, with free access to PrEP provided to all partners during ATI studies.

Several mechanisms are postulated to explain bNAb-mediated ART-free viral control. Firstly, a direct antiviral effect of bNAbs associated with serum LS-bNAb above 10 μg/mL threshold has been reported. The estimated therapeutic threshold of LS-bNAb antiviral activity remains uncertain with more recent studies ^16,18^ demonstrating a range of cut-off levels between 10-30 μg/mL. Our findings demonstrate LS-bNAb-mediated viral control in the majority of participants in Arm-A. However, we observed a wide range of LS-bNAb levels at viral rebound, suggesting that the initial LS-bNAb mediated antiviral effect may vary depending on viral sensitivity to antibodies, and that the true threshold for therapeutic serum bNAb concentrations is likely higher for most individuals than the previously reported 10 μg/mL. Possible other host factors may include autologous antibodies, innate immunity, T-cell function, genetic, epigenetic, and HIV-specific cellular effector functions as well as characteristics of the HIV reservoir.

Despite pre-screening for LS-bNAb sensitivity using genotypic methods, at least 8 participants who received LS-bNAb in Arm-A experienced viral rebound by 20 weeks at levels of serum LS-bNAb much higher than 10ug/mL, which raises the possibility of undetected or de-novo LS-bNAb resistance. We report on viral susceptibility to bNAbs of the rebound variants separately ^28^, but there was evidence for emerging resistance, which may have been pre-existing or evolved after dosing. Current algorithms used to predict LS-bNAb sensitivity remain problematic; neither *ex-vivo* phenotypic neutralisation^29^ or viral envelope sequencing^30,31^ can accurately predict clinical sensitivity^16,18,32^.

For the majority of Arm-A participants who eventually experienced HIV rebound despite prolonged viral suppression following ART interruption, viral rebound occurred as LS-bNAb plasma concentrations approached 10 μg/mL. We observed three times as many participants in Arm-A (n=7) compared with Arm-B (n=2) who maintained viral control beyond 96 weeks, a time-point at which serum bNAb concentrations are expected to have washed out, although the actual concentration in blood or tissue where bNAbs are no longer effective has not yet been determined. These findings suggest the presence of a post-bNAb viral control mechanism. The mechanisms conferring this sustained post-bNAb control remain uncertain and are of considerable interest. bNAbs have been shown to enhance pre-existing immune responses and to induce and sustain novel HIV-specific CD4 and CD8 T-cell responses^33^ in both non-human primate^34^ and human studies^35–37^, suggesting that immune mediated mechanisms may underlie prolonged viral control following bNAb exposure. Altaf et al.^38^ demonstrate a significant impact of bNAbs and ATI on HIV-specific T cell function and an association with T cell function and the duration of post-bNAb control. Fumagalli et al.^28^ report a significant correlation between baseline reservoir sensitivity to autologous antibodies to 10-1074 and time to rebound in RIO participants. In addition, a more rapid decay of intact proviral half-lives (0.65-year) compared with defective reservoir half-lifes (8.6 years) was observed following bNAbs administration^28^. Further work is ongoing to define the exact bNAb-induced immunological correlates of viral control. Finally, these findings are consistent with previously reported bNAb-mediated reductions in the intact HIV proviral reservoirs^18^, and may contribute to the observed post-ART viral control in RIO. HIV-reservoir analysis is ongoing in RIO.

There are limitations to the generalisability of our findings. Study participation was restricted to individuals who initiated ART during early-stage HIV infection, a group characterised by smaller, more homogeneous HIV reservoirs and better preserved immune responses compared with those who started ART later. The majority of participants were white cis-gender men, with clade B virus, reflecting the UK demographics of people diagnosed during early-stage HIV. However, a study of CAP256V2-LS and VRC07-523LS with vesatolimod in African women, with primarily subtype C virus, reported similar results, with 6 of 20 participants experiencing delayed viral rebound within 48 weeks^39^.

Maintenance of viral load below the transmission threshold, with duration of remission greater than 2 years, has been proposed as a minimum target product profile for a cure or remission agent for HIV ^41^. We observed seven participants with prolonged viral control past 96 weeks after receiving LS-bNAbs, with presumed sub-therapeutic serum concentrations. This was higher than the expected frequency of post-treatment control (PTC) beyond 96 weeks seen in Arm-B and frequencies of 6% in early treated people living with HIV reported in a recently published meta-analysis^6^. Some participants had detectable VL above 200 copies/mL but less than 1000 copies/mL. Within routine clinical practice, a plasma HIV VL >200 copies/mL would prompt modification of ART. However, in this scenario, where we aimed to limit viral rebound, a more relaxed criteria of 1000 copies/ml was used. The ultimate goal is to identify interventions which maintain viral control below the recognised transmission threshold^40^ in the absence of ART, as a step towards HIV remission. Future HIV cure trial strategies may include LS-bNAbs in combination with other small molecules^41^ or immunomodulatory agents to enhance HIV-specific cellular responses. These include therapeutic vaccinations, reversal of immune checkpoint blockade (e.g. anti-PD1, IL-15), with small proof-of-concept studies^42,43^ and non-human primate studies^44^ supporting their use.

RIO demonstrates the longest reported periods of ART-free viral remission following two LS-bNAbs or any other immunotherapeutic intervention in a prospective, double-blind randomised controlled trial. The study team would like to acknowledge the extraordinary commitment of all RIO participants, their partners and families. Three-quarters of participants remained off ART for 20 weeks after a single dose of the LS-bNAbs, and ART-free viral control beyond 96 weeks was more frequent in the bNAb group than in the placebo group, despite presumed sub-therapeutic serum concentrations. LS-bNAbs represent a promising step towards achieving ART-free HIV control and remission, with sustained community engagement underscoring the importance of continued cure-focused research.

## Data Availability

All data produced in the present study are available upon reasonable request to the authors

## Acknowledgments and funders

We gratefully acknowledge all participants and the clinical staff at each site involved in the RIO Trial. The complete list of staff is available in the supplementary materials. RIO Trial was funded by The Gates Foundation (OPP1210792). S.F., and H.B., and the Imperial Clinical Trials Unit acknowledge support from the NIHR Imperial Biomedical Research Centre (BRC). MCN and MC acknowledge support from the Stavros Niarkos Foundation and Robert Wennett. MCN is a Howard Hughes Medical Institute (HHMI) Investigator. Infrastructure support for this research was provided by the NIHR Imperial Biomedical Research Centre (BRC) and the NIHR Imperial Clinical Research Facility (CRF). MJL received funding from the Medical Research Council UK for a Clinical Research Training Fellowship (MR/W024454/1). JF acknowledges support from the NIHR Oxford Biomedical Research Centre and the Martin Delaney REACH Collaboratory. KES and GDT acknowledge funding by the Gates Foundation (INV-036842) and the NIH Center for AIDS Research (P30AI064518). The remaining authors do not have any competing interests to declare. The funders were not involved in the study desigh, data collection, analysis, or interpretation of data, the writing of this article, or the decision to submit it for publication.

## Data sharing statement

Individual de-identified participant data collected during the trial (including data dictionaries) will be available to share, on application, once the trial has completed and the database has been locked. Study documents including the protocol, statistical analysis plan (SAP) will be available on request and in this publication.

Proposals/requests should be directed to the Chief Investigator and requestors will be asked to sign a data access agreement. Data sharing requests will be shared with IDMC and TSC for their review and approval.

## Author contributions

SF is the chief investigator. JF is the laboratory lead and co-chief investigator. MCN and MC are the co-investigators at Rockefeller University. MJL was the trial physicians at Imperial College London and was responsible for preparation of the first draft of the manuscript. TE is the subsequent trial physician at Imperial College London. L-RC was responsible for the unblinded statistical analyses, as defined in the SAP. PZ was responsible for the bNAb screening for study eligibility determination. L-RC and EF are the trial statisticians. SC is the community representative for people living with HIV. HB and SF are responsible for the overall management, coordination, governance, and conduct of the study. HBr, NR, TT, MF, CB, AK, KS, GT, MEA, JAW, are responsible for HIV immunology, viral laboratory endpoints, bNAb and ADA measurement assays. GPT, IJ and JU are responsible for the central laboratory processing for the samples from UK participants. MJL, SF, JFo, AU, AC, SK-dL, SP, MB, GW, OSS, JDG, KR, IM, JG, PG, HN, LH, RS, PC, and CO are clinical investigators responsible for recruitment and participant care. JH, MCN, and MC designed and developed the investigational medicinal products and contributed to the study design. All authors contributed to the writing and review of the manuscript. Authorship eligibility for this manuscript and all future manuscript output related to this study will be in accordance with the International Committee of Medical Journal Editors criteria for authorship (http://www.icmje.org/recommendations/browse/roles-and-responsibilities/defining-the-role-of-authors-and-contributors.htmL). No professional writers were involved in this work or are intended to be involved in future manuscripts. All staff involved in the study are listed in the supplementary appendix.

This article is subject to HHMI’s Open Access to Publications policy. HHMI lab heads have previously granted a non-exclusive CC BY 4.0 license to the public and a sublicensable license to HHMI in their research articles. Pursuant to those licenses, the author-accepted manuscript of this article can be made freely available under a CC BY 4.0 license immediately upon publication.

