## Supplementary Material for "Time to HIV rebound after antiretroviral therapy interruption: a double-blind randomised placebo-controlled trial of long-acting broadly neutralising antibodies; The RIO Trial"

**Supplementary Figures**

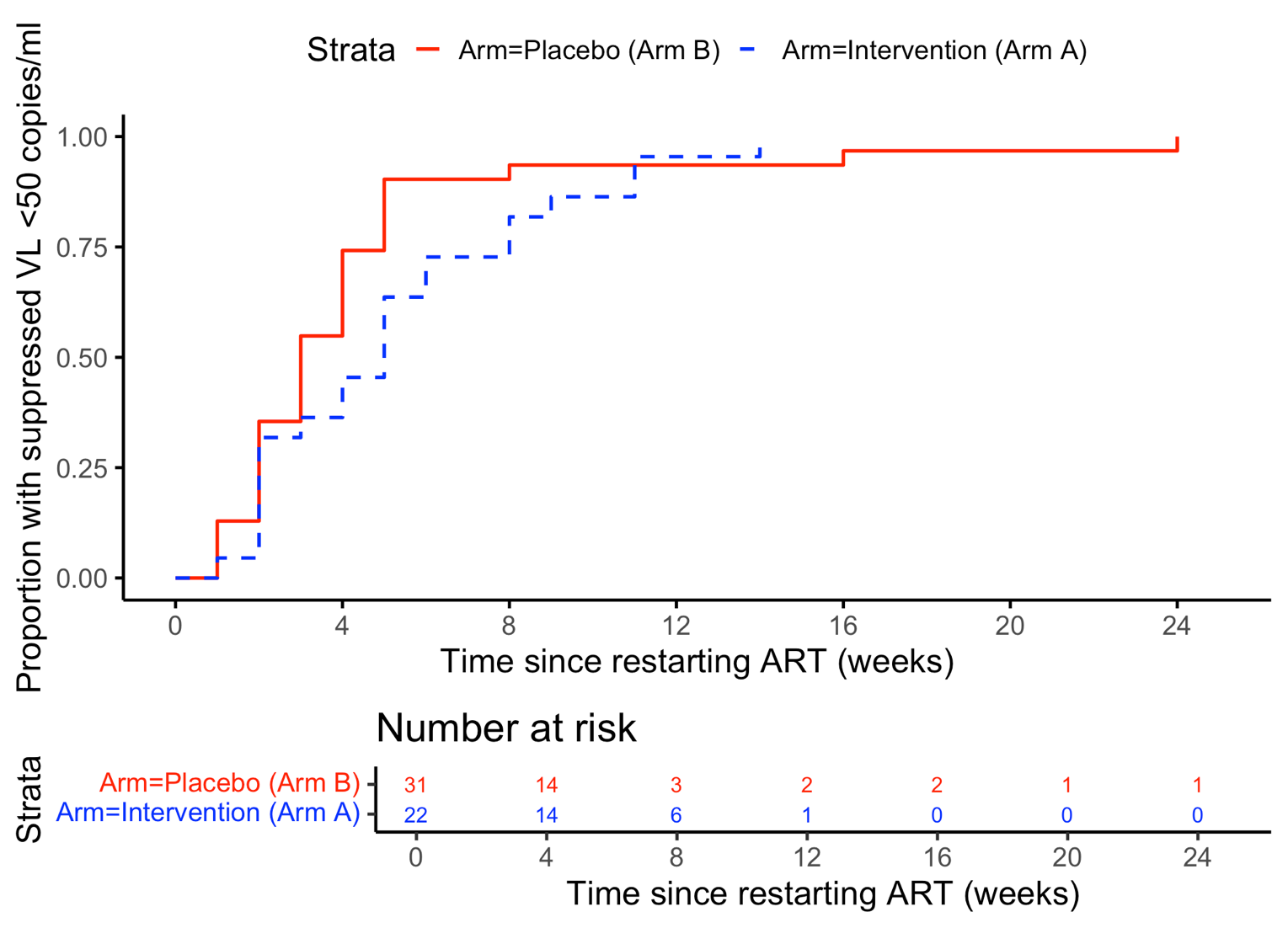
**Figure S1**

Kaplan-Meier (KM) survival curves showing the proportion of participants who experienced viral resuppression following ART restart after completion of ATI, stratified by study arm (dashed = placebo (arm-B), solid = bNAbs (arm-A)

**Table S1. List of clinical sites and HIV viral load assay platforms**

| Site | HIV Viral Load assay | Limit  (Copies/mL) |
| --- | --- | --- |
| 001 – Imperial College Healthcare NHS Trust | Roche-Cobas  5800/6800/8800 | <20 |
| 002 – Royal Free Hospital NHS Foundation Trust | Aptima™ HIV-1 Quant Dx | <40 |
| 003 – Guys and St Thomas Hospital NHS Foundation Trust | Abbott Alinity M | <20 |
| 004 – Barts Health NHS Trust | Roche-Cobas 5800/6800/8800 | <50 |
| 005 – Central and North West London NHS Foundation Trust | Aptima™ HIV-1 Quant Dx | <50 |
| 006 – Chelsea and Westminster Hospital NHS Foundation Trust | Roche- Cobas  5800/6800/8800 | <20 |
| 007 – University Hospital Sussex NHS Foundation Trust | Abbott Real Time HIV-1 assay | <40 |
| 008 – Manchester University NHS Foundation Trust | Roche-Cobas 5800/6800/8800 | <50 |
| 010 – Western General Hospital, NHS Lothian Trust | Abbot Alinity M | <20 |
| 012 – Aarhus University and Aarhus University Hospital | Roche-Cobas 5800/6800/8800 | <20 |

**Table S2. Anti-drug antibodies results**

| Participant | Number of bNAb doses | Baseline  pre-bNAb | 12 weeks post 1^st^ dose | 20 - 24 weeks post 1^st^ dose  (Pre 2^nd^ dose) | 12 weeks post 2^nd^ dose | 24 weeks post 2^nd^ dose |
| --- | --- | --- | --- | --- | --- | --- |
| 10-1074-LS | | | | | | |
| 1 | 1 | N | N | N | - | - |
| 2 | 1 | N | N | N | - | - |
| 3 | 1 | N | N | N | - | - |
| 4 | 2 | N | N | N | N | N |
| 5 | 2 | N | N | N | N | N |
| 6 | 2 | N | N | N | N | N |
| 7 | 2 | N | N | N | N | N |
| 8 | 2 | N | N | N | N | N |
| 9 | 2 | N | N | N | - | - |
| 10 | 2 | N | N | N | N | N |
| 11 | 2 | N | N | - | N | N |
| 3BNC117-LS | | | | | | |
| 1 | 1 | N | N | N | - | - |
| 2 | 1 | **P** | N | N | - | - |
| 3 | 1 | N | N | N | - | - |
| 4 | 2 | N | N | N | N | N |
| 5 | 2 | N | N | N | N | N |
| 6 | 2 | N | N | N | N | N |
| 7 | 2 | N | N | N | N | N |
| 8 | 2 | N | N | N | N | N |
| 9 | 2 | N | N | N | - | - |
| 10 | 2 | N | N | N | N | N |
| 11 | 2 | N | N | - | N | N |

Results of two-tiered confirmatory anti-drug-antibody (ADA) assays to 10-1074-LS and 3BNC117-LS. N = Negative for ADA. P = Positive for ADA. (-) = Sample not available. One participant had evidence of treatment independent ADA pre-bNAb dosing, at low titer, and none at follow-up time points after bNAb dosing.

**Table S2. Individual details of severe adverse events and related adverse events**

| Number | Arm | Class | Description | Duration | Severity | Outcome | Relation |
| --- | --- | --- | --- | --- | --- | --- | --- |
| **Severe Adverse Events** | | | | | | | |
| 005-009 | bNAb | Resulted in Death | Myocardial infarction | 1 days | Grade 5 (Fatal) | Fatal | Not Related |
| 001-032 | bNAb | Hospitalisation | Enterovirus infection | 2 days | Grade 3 (Severe) | Resolved | Unlikely |
| 003-005 | Placebo | Life-threatening | Attempted suicide | 1 days | Grade 3 (Severe) | Resolved | Not Related |
| 006-010 | Placebo | Hospitalisation | Perianal abscess | 2 days | Grade 3 (Severe) | Resolved | Unlikely |
| 006-014 | Placebo | Hospitalisation | Elbow cellulitis | 7 days | Grade 2 (Moderate) | Resolved | Unlikely |
| 007-002 | Placebo | Hospitalisation | Chronic cystitis | 4 days | Grade 3 (Severe) | Resolved with Sequelae | Not Related |
| 007-002 | Placebo | Hospitalisation | Obstructive uropathy | 2 days | Grade 2 (Moderate) | Resolved with Sequelae | Not Related |
| 007-002 | Placebo | Hospitalisation | Pyelonephritis | 8 days | Grade 2 (Moderate) | Resolved | Not Related |
| 007-002 | Placebo | Hospitalisation | Epididymo-orchitis | 6 days | Grade 2 (Moderate) | Resolved with Sequelae | Not Related |
| **Related Adverse Events** | | | | | | | |
| **Arm A (LS-b Nabs)** | | | | | | | |
| 1 | 2 | bNAb | raised ALT | Grade 1 (Mild) | Possibly | Not Related | Resolved |
| 2 | 4 | bNAb | Fatigue | Grade 1 (Mild) | Possibly | Possibly | Resolved |
| 3 | 9 | bNAb | Enterovirus infection | Grade 3 (Severe) | Unlikely | Possibly | Resolved |
| 4 | 12 | bNAb | Diarrhoea | Grade 1 (Mild) | Possibly | Not Related | Resolved |
| 5 | 15 | bNAb | Chest tightness | Grade 1 (Mild) | Possibly | Possibly | Resolved |
| 6 | 15 | bNAb | Sore throat | Grade 1 (Mild) | Possibly | Possibly | Resolved |
| 7 | 18 | bNAb | Light headedness day after 2nd dose | Grade 1 (Mild) | Possibly | Possibly | Resolved |
| 8 | 18 | bNAb | Anaemia | Grade 1 (Mild) | Possibly | Possibly | Persisting |
| 9 | 19 | bNAb | Sore arm from cannula | Grade 1 (Mild) | Possibly | Not Related | Resolved |
| 10 | 19 | bNAb | Dizziness following venepuncture (250ml blood drawn) | Grade 1 (Mild) | Not Related | Definitely | Resolved |
| 11 | 20 | bNAb | Cough | Grade 2 (Moderate) | Possibly | Not Related | Resolved |
| 12 | 20 | bNAb | Bruising | Grade 1 (Mild) | Not Related | Definitely | Resolved |
| 13 | 25 | bNAb | Covid-19 | Grade 2 (Moderate) | Not Related | Possibly | Resolved |
| 14 | 27 | bNAb | Decreased CD4:CD8 ratio | Grade 1 (Mild) | Not Related | Definitely | Resolved with Sequelae |
| 15 | 27 | bNAb | Decreased CD4/CD8 ratio | Grade 1 (Mild) | Not Related | Definitely | Resolved |
| 16 | 28 | bNAb | Left arm pain during big bleed | Grade 1 (Mild) | Not Related | Possibly | Resolved |
| 17 | 28 | bNAb | Episode of dizziness | Grade 1 (Mild) | Not Related | Possibly | Resolved |
| 18 | 29 | bNAb | vasovagel episode | Grade 1 (Mild) | Unlikely | Possibly | Resolved |
| 19 | 31 | bNAb | Fatigue | Grade 1 (Mild) | Possibly | Possibly | Resolved |
| **Arm B (Placebo)** | | | | | | | |
| 1 | 1 | Placebo | Headache | Grade 1 (Mild) | Possibly | Unlikely | Resolved |
| 2 | 1 | Placebo | Fatigue | Grade 1 (Mild) | Possibly | Not Related | Resolved |
| 3 | 3 | Placebo | 'Heavy headed' for about 20 seconds during 10-1074-LS infusion | Grade 1 (Mild) | Possibly | Possibly | Resolved |
| 4 | 3 | Placebo | Neutropenia | Grade 2 (Moderate) | Not Related | Possibly | Resolved |
| 5 | 5 | Placebo | New antiretroviral resistance-associated mutation | Grade 2 (Moderate) | Not Related | Possibly | Resolved with Sequelae |
| 6 | 5 | Placebo | Lethargy | Grade 1 (Mild) | Possibly | Possibly | Resolved |
| 7 | 5 | Placebo | lethargy | Grade 1 (Mild) | Probably | Possibly | Resolved |
| 8 | 5 | Placebo | Presyncope | Grade 2 (Moderate) | Not Related | Definitely | Resolved |
| 9 | 6 | Placebo | Diarrhoea | Grade 1 (Mild) | Possibly | Unlikely | Resolved |
| 10 | 6 | Placebo | Lethargy | Grade 1 (Mild) | Possibly | Possibly | Resolved |
| 11 | 6 | Placebo | Insomnia | Grade 1 (Mild) | Unlikely | Possibly | Resolved |
| 12 | 6 | Placebo | Fatigue | Grade 1 (Mild) | Possibly | Possibly | Resolved |
| 13 | 7 | Placebo | Somnolence | Grade 1 (Mild) | Possibly | Possibly | Resolved |
| 14 | 8 | Placebo | Lethargy | Grade 1 (Mild) | Possibly | Possibly | Resolved |
| 15 | 10 | Placebo | COVID-19 | Grade 2 (Moderate) | Not Related | Possibly | Resolved |
| 16 | 11 | Placebo | Hot flush | Grade 1 (Mild) | Possibly | Probably | Resolved |
| 17 | 13 | Placebo | Vasovagal during big bleed study procedure | Grade 2 (Moderate) | Not Related | Definitely | Resolved |
| 18 | 14 | Placebo | Dizzyness (following big bleed) | Grade 1 (Mild) | Not Related | Definitely | Resolved |
| 19 | 16 | Placebo | Fatigue | Grade 1 (Mild) | Possibly | Possibly | Resolved |
| 20 | 17 | Placebo | L. arm pain at cannula site post-infusion | Grade 1 (Mild) | Definitely | Not Related | Resolved |
| 21 | 21 | Placebo | Tired | Grade 1 (Mild) | Possibly | Possibly | Resolved |
| 22 | 21 | Placebo | Headache | Grade 1 (Mild) | Possibly | Possibly | Resolved |
| 23 | 21 | Placebo | Fatigue | Grade 2 (Moderate) | Possibly | Not Related | Resolved |
| 24 | 22 | Placebo | Bruising ACF (antecubital fossa) | Grade 1 (Mild) | Not Related | Definitely | Resolved |
| 25 | 22 | Placebo | Nausea | Grade 1 (Mild) | Possibly | Possibly | Resolved |
| 26 | 23 | Placebo | Perianal Abscess | Grade 3 (Severe) | Not Related | Possibly | Resolved |
| 27 | 24 | Placebo | Skin rash | Grade 1 (Mild) | Possibly | Not Related | Persisting |
| 28 | 26 | Placebo | Flatulence | Grade 1 (Mild) | Possibly | Unlikely | Persisting |
| 29 | 30 | Placebo | Bruises left and right arm following cannulation | Grade 1 (Mild) | Not Related | Possibly | Resolved |
| 30 | 32 | Placebo | Fatigue | Grade 1 (Mild) | Not Related | Possibly | Resolved |

**Table S3. Population parameters and 90% prediction intervals**

| Population Parameters | 10-1047LS | 90% Pred Intervals | 3BNC117LS | 90% Pred Intervals |
| --- | --- | --- | --- | --- |
| A_pop | 4.00 x 10^-15^ | 3.99 x10^-15^ – 4.01 x10^-15^ | 7.86 x 10^-5^ | 7.86 x 10^-5^ – 7.86 x 10^-5^ |
| B_pop | 2.51 x 10^-4^ | 2.38 x10^-4^ – 2.63 x10^-4^ | 1.19 x 10^-4^ | 1.14 x 10^-4^ – 1.28 x 10^-4^ |
| ɑ_pop | 1.01 x 10^9^ | 9.78 x10^8^ – 1.01 x10^8^ | 5.23 x 10^-2^ | 5.23 x 10^-2^ - 5.23 x 10^-2^ |
| ᵝ_pop | 9.65 x10^-3^ | 9.09 x10^-3^ – 1.00 x10^-2^ | 1.07 x 10^-2^ | 1.04 x 10^-2^ – 1.12 x 10^-2^ |
| ᵝ_B_logWt | 0.00 | -0.91 – 0.00 | 0.00 | -1.10 – 0.00 |
| Ω_A | 3.07 x10^-8^ | 2.30 x10^-8^ – 1.05 x10^-5^ | 1.71 x 10^-8^ | 2.47 x 10^-8^ – 3.08 x 10^-8^ |
| Ω_B | 1.86 x10^-1^ | 5.37 x10^-8^ – 2.00 x10^-1^ | 1.12 x 10^-1^ | 7.51 x 10^-2^ – 1.55 x 10^-1^ |
| Ω_ɑ | 6.68 x10^-8^ | 2.50 x10^-8 –^ 5.58 x10^-3^ | 6.84 x 10^-8^ | 2.23 x 10^-8^ – 1.87 x 10^-7^ |
| Ω_ᵝ | 1.55 x10^-1^ | 8.94 x10^-2^ – 1.96 x10^-1^ | 2.29 x 10^-1^ | 1.19 x 10^-1^ – 2.59 x 10^-1^ |
| Elimination half life | 72.5 days | 69.1 – 76.2 days | 64.8 days | 61.8 – 66.6 days |

**Table S4. CD4, CD8, and CD4/CD8 ratios**

| Arm | Visit | N | Mean | SD | Median | Min | Max |
| --- | --- | --- | --- | --- | --- | --- | --- |
| **CD4 counts** | | | | | | | |
| Placebo | Screening | 34 | 801 | 280 | 810 | 272 | 1,352 |
| Placebo | Week 12 | 5 | 773 | 111 | 727 | 666 | 920 |
| Placebo | Week 20 | 4 | 736 | 115 | 717 | 639 | 870 |
| Placebo | 12 weeks post 2nd dose | 4 | 675 | 150 | 674 | 501 | 850 |
| Placebo | 24 weeks post 2nd dose | 3 | 681 | 253 | 584 | 490 | 968 |
| bNAb | Screening | 34 | 799 | 210 | 742 | 511 | 1,468 |
| bNAb | Week 12 | 30 | 808 | 260 | 749 | 413 | 1,487 |
| bNAb | Week 20 | 22 | 805 | 283 | 743 | 442 | 1,550 |
| bNAb | Week 32 (no 2nd dose) | 3 | 1,019 | 362 | 987 | 675 | 1,396 |
| bNAb | 12 weeks post 2nd dose | 14 | 835 | 323 | 712 | 523 | 1,638 |
| bNAb | 24 weeks post 2nd dose | 13 | 835 | 369 | 677 | 540 | 1,869 |
| **CD8 counts** | | | | | | | |
| Placebo | Screening | 34 | 670 | 292 | 717 | 168 | 1,398 |
| Placebo | Week 12 | 5 | 721 | 270 | 840 | 270 | 913 |
| Placebo | Week 20 | 4 | 582 | 255 | 606 | 250 | 867 |
| Placebo | 12 weeks post 2nd dose | 4 | 529 | 99 | 497 | 453 | 670 |
| Placebo | 24 weeks post 2nd dose | 3 | 665 | 73 | 696 | 582 | 717 |
| bNAb | Screening | 34 | 704 | 215 | 674 | 383 | 1,279 |
| bNAb | Week 12 | 30 | 720 | 236 | 698 | 379 | 1,142 |
| bNAb | Week 20 | 22 | 746 | 234 | 766 | 384 | 1,114 |
| bNAb | Week 32 (no 2nd dose) | 3 | 845 | 391 | 755 | 506 | 1,273 |
| bNAb | 12 weeks post 2nd dose | 14 | 753 | 221 | 730 | 479 | 1,252 |
| bNAb | 24 weeks post 2nd dose | 13 | 734 | 259 | 687 | 360 | 1,246 |
| **CD4/CD8 ratio** | | | | | | | |
| Placebo | Screening | 34 | 1.37 | 0.59 | 1.25 | 0.78 | 3.48 |
| Placebo | Week 12 | 5 | 1.29 | 0.74 | 1.10 | 0.73 | 2.56 |
| Placebo | Week 20 | 4 | 1.51 | 0.76 | 1.38 | 0.74 | 2.56 |
| Placebo | 12 weeks post 2nd dose | 4 | 1.29 | 0.25 | 1.32 | 0.96 | 1.55 |
| Placebo | 24 weeks post 2nd dose | 3 | 1.01 | 0.33 | 0.84 | 0.81 | 1.39 |
| bNAb | Screening | 34 | 1.21 | 0.43 | 1.16 | 0.63 | 2.54 |
| bNAb | Week 12 | 30 | 1.19 | 0.39 | 1.15 | 0.60 | 2.36 |
| bNAb | Week 20 | 22 | 1.13 | 0.34 | 1.08 | 0.57 | 1.75 |
| bNAb | Week 32 (no 2nd dose) | 3 | 1.25 | 0.13 | 1.31 | 1.10 | 1.33 |
| bNAb | 12 weeks post 2nd dose | 14 | 1.16 | 0.43 | 1.02 | 0.68 | 1.81 |
| bNAb | 24 weeks post 2nd dose | 13 | 1.21 | 0.45 | 1.04 | 0.54 | 1.87 |

Supplementary methods:

**HIV envelope amplification and sequencing methods**

**Nested Envelope PCR amplification.** Extracted DNA were amplified using a nested envelope PCR protocol, with one of three primer sets: Default protocol (Outer reaction primers: Env_B5Out, Env_B3Out; Inner reaction protocol: Env_B5In, Env_B3In), Long protocol (LP) (Outer reaction primers: 5’_B3R3, 3’_R3B6R; Inner reaction protocol: Env_B5In, Env_B3In), and the LP807 protocol (Outer reaction primers: 5’_B3R3, 3’_R3B6R; Inner reaction protocol: Env_B5In, Env 807). The list of primers and thermocycler protocols are listed in Table S6.

Single-genome amplification (SGA) of the HIV env gene was performed to ensure amplification from individual viral templates. To achieve this, sample dilutions were optimized so that no more than 30% of PCR reactions yielded a product of the expected size. This approach confers a ~85% probability that each positive reaction originates from a single viral genome, minimizing the risk of in vitro recombination and resampling.. A master mix was made up of the following: For the default outer reaction and all inner reactions, a 14.25µL mix per well was made up of 11.5125µL Molecular-grade DNase/RNase-free water, 1.5µL 10X PCR buffer, 0.6µL 50mM DNA-free Magnesium Chloride, 0.0375µL DNA-free Invitrogen™ Platinum™ Taq (Fisher Scientific), 0.3µL deoxynucleotide triphosphate (dNTP) mix (Fisher Scientific, 10mM each), 0.15µL each of the forward and reverse primer. 1µL of the DNA sample was added as the template to the mix per well, for a total reaction volume of 15.25µL per well. For the LP and LP807 outer reaction reactions, a 19µL mix per well was made up of 15.3µL Molecular-grade DNase/RNase-free water, 2µL 10X PCR buffer, 0.8µL 50mM DNA-free Magnesium Chloride, 0.1 µL DNA-free Invitrogen™ Platinum™ Taq (Fisher Scientific), 0.4µL deoxynucleotide triphosphate (dNTP) mix (Fisher Scientific, 10mM each), 0.2µL each of the forward and reverse primer. 1µL of the DNA sample was added as the template to the mix per well, for a total reaction volume of 20µL per well. After amplification of the inner reaction, the samples were diluted five-fold, and underwent DNA electrophoresis on a pre-cast 96well 1% Invitrogen™ agarose E-gel (Fisher Scientific) for 12minutes on an E-base™ (Fisher Scientific) under program EG as per manufacturer’s instructions, with an E-gel sizing DNA ladder marker (Fisher Scientific). If >30% of wells were positive for appropriately sized Env PCR products (~3000bp), the experiment was repeated using an optimised sample dilution to achieve the target. Amplicons from positive SGA reactions were used for DNA library preparation described in the next step. A target of twenty single genome sequences were obtained per participant.

**DNA Library preparation.** DNA library preparation is performed using a custom library preparation protocol. The amplified Env products are diluted three-fold with Molecular-grade DNase/RNase-free water. For the tagmentation of samples from a 96-well plate, a tagmentation master mix comprising of 132µL of Tagmentation buffer and 26.4µL of tagmentation enzyme (TDE-1 Tagment DNA Enzyme and Buffer kit, Illumina®) was mixed and 1.5µL of the mix was distributed to each well of the reaction plate. 1µL of the PCR product was added into each well and incubated at 55^o^C for 5 minutes. Barcode index adapters (8x i5 and 12x i7, Nextera XT Index Kit v2 Set A, Illumina®) were prepared as index master mixes by adding 18µL of 2X KAPA HiFi HS Ready Mix polymerase (Roche) and 6µL of i7 index adapters, and 27µL of 2X KAPA polymerase and 9µL of i5 index adapters respectively. 2.5µL of each i5 and i7 index adapter master mix were added to each well using a multichannel pipette resulting in 96 individual permutation of the two adapters per well for individual barcoding. The reaction plate was then placed in the thermocycler to run the tagmentation protocol listed in Table 2. Following tagmentation, the resultant product was cleaned up using Ampure XP beads (Beckman Coulter™). 8 wells of tagmentation product were pooled in a 1.5mL Eppendorf tube and 136µL of Ampure XP beads added and incubated at RT for 5 minutes. The Eppendorf tubes were then placed on a magnetic stand and incubated for 1 minute, followed by removal of the supernatant without disturbing the beads on the side of the tube. The beads were washed with 70% ethanol 4 times and left to air-dry after removing the last ethanol wash step. After removing the Eppendorf tubes from the magnetic stand, the beads were resuspended in 100µL of Tris-EDTA (TE) buffer and incubated for a further 2 minutes. The Eppendorf tubes were placed on the magnetic stand again, and the supernatant was collected from each tube and pooled into a single DNA Lobind Eppendorf tube for the DNA library.

**Table S6. List of HIV Envelope nested single genome amplification primers and thermocycler protocols**

| List of primers | | | |
| --- | --- | --- | --- |
| Target region | Primer name | Sequence (5’ – 3’) | Position (HXB ref) |
| Env | Env_B5Out | TAGAGCCCTGGAAGCATCCAGGAAG | 5853 – 5877 |
|  | Env_B3Out | TTGCTACTTGTGATTGCTCCATGT | 8913 – 8936 |
|  | 5’_B3R3 | TGGAAAGGTGAAGGGGCAGTAGTAATAC |  |
|  | 3’_R3B6R | TGAAGCACTCAAGGCAAGCTTTATTGAGGC |  |
|  | Env_B5In | CACCTTAGGCATCTCCTATGGCAGGAAGAAG | 5953 – 5983 |
|  | Env_B3In | GTCTCGAGATACTCGTCCCACCC | 8882 – 8904 |
|  | Env_807 | GTCTCGAGACGCTGGTCCTACTC | 8882 – 8904 |
| Thermocycler protocols | | |  |
|  | Temperature | Time | No. of cycles |
| Env Default  Outer Protocol | 94 ^o^C | 2 minutes | 1x |
|  | 94 ^o^C | 15 seconds | 35x |
|  | 58.5^o^C | 30 seconds |  |
|  | 68 ^o^C | 4 minutes |  |
|  | 68 ^o^C | 15 minutes | 1x |
|  | 4 ^o^C | For storage |  |
| Env Long Outer Protocol | 94 ^o^C | 2 minutes | 1x |
|  | 94 ^o^C | 15 seconds | 45x |
|  | 55^o^C | 30 seconds |  |
|  | 68 ^o^C | 4 minutes |  |
|  | 68 ^o^C | 15 minutes | 1x |
|  | 4 ^o^C | For storage |  |
| Env Inner Protocol | 94 ^o^C | 2 minutes | 1x |
|  | 94 ^o^C | 15 seconds | 44x |
|  | 61^o^C | 30 seconds |  |
|  | 68 ^o^C | 4 minutes |  |
|  | 68 ^o^C | 15 minutes | 1x |
|  | 4 ^o^C | For storage |  |
| Tagmentation protocol | 72 ^o^C | 3 minutes | 1x |
|  | 98 ^o^C | 2minutes 45 seconds | 1x |
|  | 98^o^C | 15 seconds | 8x |
|  | 62 ^o^C | 30 seconds |  |
|  | 72 ^o^C | 1 minute 30 seconds |  |
|  | 4 ^o^C | For storage |  |

**Sequencing**

The DNA library was sequenced using the MiSeq System (Illumina®), and MiSeq Reagent Kits v3 (Illumina®) at the Frater lab, University of Oxford. The output was delivered in demultiplexed fastq files.

**BNAb screening**

A custom bioinformatics pipeline was employed to process HIV sequencing data on a per-sample basis previously described by West et al. (20). First, all reads not aligning to HIV were discarded. The remaining HIV-aligned reads were used to generate an initial consensus reference, selected by aligning to the most closely related HIV *env* reference sequence. Reads were then re-aligned to this initial reference to generate a refined, sample-specific final reference. A second round of alignment was performed against this final *env* reference. To ensure single-template representation, the pipeline flagged sequences with >30% base disagreement at any position, indicating potential contamination with multiple templates. Dysfunctional *env* sequences were also discarded. Finally, high-confidence *env* sequences were translated into protein for downstream analysis. BNAb resistance prediction were computed using a custom bnAb sensitivity prediction model, previously described by Gaebler et al. (16). The specific amino-acid residues associated with resistance in this algorithm are as follow: for 10-1074 325D/N, 330H/Y, 332N, 333notP, 334S/T; for 3BNC117 279D/N, 456R, 457D, 458G, 459G, and the output is a binary prediction of resistant or not resistant for each sequence. If 15% or more sequences were predicted to have resistance-associated mutations, the individual were deemed ineligible for enrolment in the RIO study.

### RIO Study Group

**Chief Investigator:** Sarah Fidler
**Protocol Co-Chair:** John Frater
**Co-Investigators:** Michel Nussenzweig, Marina Caskey

**Trial Investigators:** Sarah Fidler, John Frater, Graham Taylor, Michel Nussenzweig, Marina Caskey, Saye Khoo, Ming Lee, Simon Collins, Julie Fox, Amanda Clarke, Sabine Kinloch-de Loes, Sarah Pett, Kyle Ring, Chloe Orkin, Marta Boffito, Gary Whitlock, Rebecca Sutherland, Alison Uriel, Marcelino Molina, Louise Terry, Ole Schmeltz Søgaard, Jesper Damsgaard Gunst, Henrik Nielsen, Jesal Gohil, Tamara Elliott, Hanna Box, Stephen Fletcher, Louise-Rae Cherrill, Emanuela Falaschetti, Daphne Babalis, Christina Prechtl, Najwa Soussi, Milaana Jacob, Ambreen Ashraf, Toby Prevost, Nicholas Johnson, Mariam Habib, Marta Gielniewska, Jodi Meyerowitz, Mary Cross, Verity Leeson, Amnah Mirza, Jonathan Dao, Eloise Britten, Vivienne Okona-Mensah, Amanda Bravery, Francesco Lala, Nayan Das, Smita Das, Lee Barker, Maathini Balachandran, Katie Topping, Jacquie Ujetz, Ishrat Jahan, Andrew Lovell, Maryam Khan, Helen Brown, Nicola Robinson, Penny Zacharopoulou, Timothy Tipoe, Ane Ogbe, Matthew Pace, Mohammed Altaf, Marcilio Fumagalli, Anna Kaczynska, Cintia Bittar

**Community Representatives:** Simon Collins, Ben Cromarty, Jo Josh, Roy Trevelion

**Trial Steering Committee (TSC):** Frank Post (Independent Chair), Caroline Sabin, Clifford Leen, Jaime Vera, Dimitra Peppa, Ben Cromarty

**Independent Data Monitoring Committee (IDMC):** Abdel Babiker (Independent Chair), Jane Anderson, Andrew Lever, Jo Josh, Roy Trevelion (previous member)

**Endpoint Adjudication Committee (EAC):** Linos Vandekerckhove (Independent Chair), Beatriz Mothe Pujadas, Casper Rokx, Ole Schmeltz Søgaard (previous Chair)

**Investigators and Staff at Participating Sites:**

**Sarah Fidler**, Ming Lee, Marcelino Molina, Jesal Gohil, Tamara Elliott, Euan Sutherland, Wilbert Ayap, Claire Petersen, Clive Matthews, Ian McGuinness, Charlotte Blake, Sophie Taylor, Romina Tajik, Rebecca Hall, Zoe Wiggins. **Sabine Kinloch-de Loes**, Jonathan Edwards, Thomas Allan, Katie Spears, Thomas Fernandez, Jia Bo He, Abigail Tobin, Fiona Burns, Megan Bailey, Mike Young, Pedro Simoes, Nnenna Ngwu, Margaret Johnson, Qayo Egeh. **Julie Fox**, Louise Terry, Piyumika Godakandaarachchi, Kathy Arbis, Alice McKain, Jacqueline O’Connell, Anele Waters, Julianne Lwanga, Harry Coleman, Isabel Laszczak, Rebekah Roberts, Mariusz Racz. **Chloe Orkin**, Kyle Ring, John Thornhill, James Hand, Helena Miras, Hafiza Rahman, Isabelle Whelan, Harriet Le Voir, Hamzah Farooq, Louise Whitton. **Sarah Pett**, Sandra Coombes, Deirdre Sally, Manik Kohli, Vasanth Naidu, Alejandro Arenas-Pinto, Felicity Aiano, Gosala Gopalakrishnan, Florence Bascombe, Joe Phillips, Ebunoluwa Taiwo, Claudia Benatar, Irfaan Maan, Marzia Fiorino, Charlotte Woodward, Jose Paredes Sosa, Rhiannon Owen, Claire Mullender, Nicola Stewart, Emmi Suonpera. **Marta Boffito,** Gary Whitlock, Rizamay Balista, Serge Fedele, Alfredo Soler Carracedo, Betsabe Rodriguez Mateos, Rosalie Housman, Roya Movahedi, Ruth Byrne, Hyun Jin Kim, Freya Toyne, Ana Milinkovic, Julie Logan, Jad Salha, Clovis Salinos, Reda Yassein, Rosa Diaz Echeverria, Patrizia Simonato, Cherry Colcol, Veronica Canuto, Krestine Electito. **Amanda Clarke,** Kiersten Simmons, Tanya Adams, Mikaela Vinick, Lisa Barbour, Caroline Cable, Andrea Terlingo, Claire Norcross, Sophie Ross, Sonia Raffe, Sophie Ross, Fionnuala Finnerty, Vittorio Trevitt, Bryony Broster, Leigh Greenland, Celia Richardson, Andrew Bexley, Sarah Kirk, Collins Iwuji, Justina Strikaite, Goabaone Diteko, Donards Kim Tanedo, Marion Campbell. **Alison Uriel,** Denise Kadiu, Milisha Gihan, Gabriella Lindergard, Claire Fox, Chloe Lord, Irvine Mangawa, Pabalelo Pule, Pamela Hackney, Jacinta Guerin, Phoebe Shellman, Serah Titus, Raiza Hussain, Sam Hey, Bini George, Yu Tang, Kevin Kuriakose, Karen Talbot, Lisa Southon, Thomas Lamb, Fahd Niaz, Alice Hendy, Jan Flaherty. **Lisa Hamzah,** Aline Lopes Fernandes, Joana Teixeira, Hong Ju, Maria Belini, Samantha Taylor, Claire Gilmartin, Alkida Bucaj, Bojana Dragovic. **Rebecca Sutherland,** David Dockrell, Juergen Haas, Susie Ferguson, Amy Shepherd, Louise Sharp, Jacqueline Henderson, Karla Berry, Anna Gordon, Alexandria Chung, Penelope Saverton, Jennifer Marshall, Anne Saunderson. **Ole Schmeltz Søgaard,** Jesper Damsgaard Gunst, Ane Søndergaard, Ida Soelberg Høj, Yordanos Yehdego, Sandra Schieber, Lene Svinth Jøhnke, Marie Juul Eli. **Henrik Nielsen,** Maria Ruwald Juhl, Kristine Toft Petersen, Rikke Thisted. **Paola Cicconi,** Vanessa Fenech, Juliana Umunna, Musaiwale Kamfose, Charlotte Wells, Mohammad Iqbal, Mehreen Datoo, Vaitehi Nageshwaran. **Karen Mosley,** Lisa Hurley, John Bervin Galang, Susanne Fagerbrink, Marcus Frederick, Christos Karathanasis, Victoria Tsui, James Fletcher, Tom Cole, Sofia Coelho, Oluwatoyin Ayanjoke, Gifty Teibowei, Ernesto Angustia, Guillermo Batan, Konstantina Pastou, Natalie Man, Aneta Gupta, Orla Mulrooney, Cyndi Cruzata, Alanood Alshamari, Zoe Gardener, Theo Farah, Peter Talbot, Maniola Tanaka, Abiola Ogunleye, Joanna Schronce, Peace Usigbe, David Owen, Janahi Visakan, Maria Renguenge, Asha Vikraman, Bridget Oduro, Roisin O’Sullivan, Andrew Ravendren, Daisy Metcalf, Nicola Lawlor, Suryapriya Thovaray Balan, Sam Jawaid, Cecilia Njenga, Nana-Marie Lemm, Mumsy Mahange, Kim Sorley, Shelley Page, Relebogile Mawasha, Derecia Adelakun-Powlette, Tamunoibim Anidima, Sukumuran Ajithkumar.

**Pharmacokinetic (PK) Assay laboratory**.

**Kelly E. Seaton**, Shad Mosher, Alex Carnacchi, Sheetal Sawant, **Georgia D. Tomaras**

**Program management**

Hongmei Gao, Kelli Greene

**Anti-Drug Antibodies Assays**

Margaret E. Ackerman, Joshua A. Weiner
